# Determinants and long-term outcomes of COVID-19 undervaccination: a cohort study of 6.8 million individuals in Lombardy, Italy

**DOI:** 10.64898/2026.05.06.26352509

**Authors:** Andrea Corbetta, Katherine M. Logan, Francesca Ieva, Emanuele Di Angelantonio

## Abstract

**Background:** Receiving fewer COVID-19 vaccine doses than recommended (“undervaccination”) may increase risks of death, severe COVID-19, and post-COVID condition. However, population-scale evidence from Italy remains limited. We aimed to characterise determinants of undervaccination in Lombardy and to quantify its association with mortality, severe COVID-19, and long COVID outcomes.

**Methods:** We conducted a population-based study including all residents of Lombardy aged ≥30 years who were alive on June 1, 2022 (n=6,836,566), and followed them until Dec 31, 2024. Vaccine deficit was defined as the difference between age-specific recommended doses (three for <60 years; four for ≥60 years) and doses received, and was modelled as a time-varying exposure. Outcomes were all-cause mortality, severe COVID-19 (hospitalisation or COVID-19-related death), and long COVID defined using symptom-based ICD codes recorded ≥1 month after infection. Determinants of undervaccination were assessed using multivariable logistic regression. Age-stratified Cox models estimated adjusted hazard ratios (HRs). Counterfactual vaccination scenarios were simulated using fitted survival models.

**Results:** On June 1, 2022, 1,668,014 individuals (24·4%) were not up to date with recommended vaccination. Undervaccination was more frequent in younger adults, women, individuals born outside Europe, rural residents, and those with high comorbidity burden. During follow-up, 265,383 deaths, 52,121 severe COVID-19 events, and 23,780 long COVID events occurred. In adults aged ≥60 years, increasing vaccine deficit was associated with progressively higher risks of mortality (HR up to 1·63) and severe COVID-19 (HR up to 2·16). Associations were weaker in younger adults. For long COVID, effect estimates were modest and sensitive to outcome definition. Simulated universal booster coverage in adults ≥60 years was associated with substantial reductions in expected deaths and severe COVID-19 events.

**Conclusion:** About one in four adults in Lombardy was undervaccinated by mid-2022. An increasing vaccine deficit was associated with a higher risk of severe COVID-19 and mortality, particularly in older adults. Sustaining booster uptake in high-risk groups remains central to mitigating the COVID-19 burden.

## Background

Italy experienced one of the earliest and most severe COVID-19 outbreaks in Europe, with Lombardy as the epicentre of the initial wave in early 2020. Excess mortality during the first months of the pandemic was unprecedented in the post-war period and led to the rapid reorganisation of healthcare delivery and public health strategy.^1^ The introduction of COVID-19 vaccines in December 2020 substantially altered the trajectory of subsequent waves, reducing hospitalisation and death across age groups. ^2–4^

As vaccination campaigns progressed, recommendations evolved in response to waning immunity and emerging variants.^5,6^ By June 1, 2022, Italian national guidance recommended three doses for adults aged 60 years or younger and four doses for adults aged 60 years or older and for clinically vulnerable individuals.^7^ Consequently, vaccination status cannot be meaningfully summarised as vaccinated versus unvaccinated. ^2–4^ A more policy-relevant metric is whether individuals are up to date with the number of doses recommended for their age group at a given time. Across Europe, vaccine uptake has been socially patterned. Large registry-based studies have shown lower coverage among younger adults, migrants, and residents of socioeconomically disadvantaged areas. ^8–11^ Italian regional analyses have reported similar disparities. ^12–14^ However, most Italian studies have focused on coverage rather than on the health consequences of being behind recommended vaccination schedules.

Previous studies have suggested that undervaccination is associated with higher risks of COVID-19 hospitalisation and death, with clear dose–response patterns across age groups. ^9,15–19^. However, in Italy, large-scale analyses remain limited, particularly beyond COVID-specific outcomes.^20–23^ We therefore conducted a regionwide study of 6·8 million adults in Lombardy. Using linked administrative health data and modelling vaccine deficit relative to age-specific recommendations as a time-varying exposure, we aimed to characterise determinants of undervaccination and quantify its association with all-cause mortality, severe COVID-19, and long COVID outcomes.

## Methods

### Study design

We conducted a population-based study using routinely collected administrative data from the Lombardy Regional Health Service. Lombardy is the most populous region in Italy, with approximately 10 million residents, and operates under a universal healthcare system in which all vaccinations, testing, hospitalisations, prescriptions, and mortality events are centrally recorded. The databases used for this study included demographic registries, COVID-19 vaccination records, SARS-CoV-2 PCR testing databases, hospital discharge records, emergency department contacts, home care and care-home registries, pharmaceutical prescription data, and mortality records. Individual-level linkage across databases was achieved using encrypted unique health identifiers within a secure regional data infrastructure. Vaccination data are captured in real time through the regional immunisation registry, and mortality data are mandatorily reported, ensuring high completeness. Data were analysed within the secure regional environment in accordance with Italian data governance regulations.

### Study population

We included all individuals aged 30-105 years who were alive and registered with the Italian National Health Service in Lombardy on June 1, 2022. This date was selected because, by then, all adults had had the opportunity to complete the primary vaccination cycle and receive at least one booster dose, in accordance with national recommendations. Restricting the cohort to individuals alive on this date ensured that exposure classification reflected the contemporaneous recommended vaccination schedule. Baseline covariates were defined using all available information recorded before June 1, 2022. Individuals were followed from June 1, 2022, until the earliest of outcome occurrence, death (for non-mortality outcomes), deregistration from the regional health system, or Dec 31, 2024. After application of the exclusion criteria, 6 836 566 (Table 1) individuals were included in the primary undervaccination analysis and 6 087 977 in the extended analysis.

**Table 1:** Frequency and percentages (within subgroups) of doses received and undervaccination at June 1 2022.

| Group | N | Zero doses group | One dose group | Two dose group | Fully vaccinated | Under-vaccinated group |
| --- | --- | --- | --- | --- | --- | --- |
| **MCS class** |  |  |  |  |  |  |
| 0 | 386 9962 (56.6%) | 415 532 (10.7%) | 50 024 (1.3%) | 563 333 (14.6%) | 2 841 073 (73.4%) | 1 028 889 (26.6%) |
| 1 | 200 0433 (29.3%) | 132 207 (6.6%) | 23 665 (1.2%) | 267 706 (13.4%) | 1 576 855 (78.8%) | 423 578 (21.2%) |
| 2 | 52 1357 (7.6%) | 37 205 (7.1%) | 6 261 (1.2%) | 59 947 (11.5%) | 417 944 (80.2%) | 103 413 (19.8%) |
| 3 | 18 9138 (2.8%) | 14 806 (7.8%) | 2 727 (1.4%) | 25 187 (13.3%) | 146 418 (77.4%) | 42 720 (22.6%) |
| 4 | 10 1098 (1.5%) | 8 968 (8.9%) | 1 564 (1.5%) | 13 842 (13.7%) | 76 724 (75.9%) | 24 374 (24.1%) |
| 5 | 15 4578 (2.3%) | 19 710 (12.8%) | 3 120 (2.0%) | 22 210 (14.4%) | 109 538 (70.9%) | 45 040 (29.1%) |
| **Continent of birth** |  |  |  |  |  |  |
| European | 6 332 588 (92.9%) | 545 494 (8.6%) | 76 412 (1.2%) | 868 563 (13.7%) | 4 860 404 (76.5%) | 1 490 469 (23.5%) |
| African | 19 4608 (2.8%) | 35 677 (18.3%) | 4 932 (2.5%) | 39 076 (20.1%) | 114 923 (59.1%) | 79 685 (40.9%) |
| American | 12 8981 (1.9%) | 19 809 (15.4%) | 2 367 (1.8%) | 20 702 (16.1%) | 86 103 (66.8%) | 42 878 (33.2%) |
| Asian | 16 0702 (2.4%) | 27 173 (16.9%) | 3 636 (2.3%) | 23 707 (14.8%) | 106 186 (66.1%) | 54 516 (33.9%) |
| Other | 1402 (0.0%) | 275 (19.6%) | 14 (1.0%) | 177 (12.6%) | 936 (66.8%) | 466 (33.2%) |
| **Sex** |  |  |  |  |  |  |
| M | 3 257 760 (47.7%) | 292 372 (9.0%) | 40 228 (1.2%) | 460 085 (14.1%) | 2 465 075 (75.7%) | 792 685 (24.3%) |
| F | 3 578 806 (52.3%) | 336 056 (9.4%) | 47 133 (1.3%) | 492 140 (13.8%) | 2 703 477 (75.5%) | 875 329 (24.5%) |
| **Domicile classification** |  |  |  |  |  |  |
| Urban | 3 746 678 (54.8%) | 356 095 (9.5%) | 49 750 (1.3%) | 520 359 (13.9%) | 2 820 485 (75.3%) | 926 204 (24.7%) |
| Suburban | 3 078 121 (45.0%) | 271 196 (8.8%) | 37 393 (1.2%) | 430 253 (14.0%) | 2 339 279 (76.0%) | 738 842 (24.0%) |
| Rural | 11 767 (0.2%) | 1 137 (9.7%) | 218 (1.9%) | 1 613 (13.7%) | 8 799 (74.8%) | 2 968 (25.2%) |
| **Age class** |  |  |  |  |  |  |
| [32,40) | 679 770 (9.9%) | 78 733 (11.6%) | 13 431 (0.2%) | 146 582 (21.6%) | 441 024 (64.9%) | 238 746 (35.1%) |
| [40,45) | 597 325 (8.7%) | 68 335 (11.4%) | 10 439 (2.0%) | 115 671 (19.4%) | 402 880 (67.4%) | 194 445 (32.6%) |
| [45,50) | 747 685 (10.9%) | 80 207 (10.7%) | 11 380 (1.5%) | 130 791 (17.5%) | 525 307 (70.3%) | 222 378 (29.7%) |
| [50,55) | 806 803 (11.8%) | 74 594 (9.2%) | 11 045 (1.4%) | 122 680 (15.2%) | 598 484 (74.2%) | 208 319 (25.8%) |
| [55,60) | 810 415 (11.9%) | 69 464 (8.6%) | 9 334 (1.2%) | 109 181 (13.5%) | 622 436 (76.8%) | 187 979 (23.2%) |
| [65,70) | 676 146 (9.9%) | 47 663 (8.2%) | 5 222 (0.9%) | 61 567 (10.6%) | 466 112 (80.3%) | 114 452 (19.7%) |
| [60,65) | 580 564 (8.5%) | 58 162 (8.6%) | 6 989 (1.0%) | 81 628 (12.1%) | 529 367 (78.3%) | 146 779 (21.7%) |
| [70,75) | 553 151 (8.1%) | 39 278 (7.1%) | 4 552 (0.8%) | 5 3456 (9.7%) | 455 865 (82.4%) | 97 286 (17.6%) |
| [75,80) | 476 563 (7.0%) | 31 181 (6.5%) | 4 141 (0.9%) | 4 4175 (9.3%) | 397 066 (83.3%) | 79 497 (16.7%) |
| [80,85) | 425 581 (6.2%) | 27 953 (6.6%) | 3 560 (0.8%) | 3 3084 (7.8%) | 360 984 (84.8%) | 64 597 (15.2%) |
| >85 | 482 563 (7.1%) | 52 858 (11.0%) | 7 268 (1.5%) | 53 410 (11.1%) | 369 027 (76.5%) | 113 536 (23.5%) |
| **Total** |  |  |  |  |  |  |
|  | 6 836 566 (100%) | 628 428 (9.2%) | 87 361 (1.3%) | 952 225 (13.9%) | 5 168 552 (75.6%) | 1 668 014 (24.4%) |

### Exposure

The exposure of interest was vaccine deficit, defined as the difference between the number of COVID-19 vaccine doses recommended for an individual’s age group and the number of doses actually received. As of June 1, 2022, national recommendations in Italy specified three doses for adults younger than 60 years and four doses for adults aged 60 years or older, including booster doses introduced to address waning immunity and emerging variants. A single dose of the Johnson & Johnson vaccine administered during the primary cycle was considered equivalent to two doses, consistent with national guidance at the time. Vaccine deficit was treated as a categorical time-varying exposure. Individuals transitioned between deficit categories on the date they received additional doses, enabling dynamic updates to vaccination status throughout follow-up. This approach reduced exposure misclassification relative to fixed baseline categorisation. All COVID-19 vaccines licensed in Italy were included: Pfizer-BioNTech (BNT162b2), Oxford-AstraZeneca (ChAdOx1), Moderna (mRNA-1273), Johnson&Johnson (Ad26.COV2.S), Covishield (ChAdOx1, recombinant), and Novavax (NVX-CoV2373).

### Outcomes

The primary outcomes were all-cause mortality, severe COVID-19, and long COVID. All-cause mortality was defined using the regional demographic registry, which records the date of death for all residents. Severe COVID-19 was defined as hospitalisation with a COVID-19 diagnosis code (ICD-10 codes U07.1, U07.2, or U09.9) recorded as the primary or secondary diagnosis, or death attributed to COVID-19. Because cause-of-death information was incomplete for a substantial proportion of deaths, we additionally classified deaths occurring within two months of a positive SARS-CoV-2 PCR test as COVID-19-related, to improve the sensitivity of severe outcome ascertainment. Long COVID was defined using symptom-based diagnostic codes consistent with the WHO clinical case definition of post-COVID condition.^24–27^ Eligible symptoms included fatigue, memory loss, altered mental status, generalised pain, abnormal gait, dyspnoea, and other related codes (ICD codes available in Appendix p25) recorded in hospital, emergency, home-care, or care-home settings. To reduce misclassification of pre-existing symptoms, only diagnoses recorded at least one month after a positive PCR test were considered. In sensitivity analyses, we removed the requirement for PCR-confirmed infection to account for possible under-testing during later pandemic phases.

## Statistical Analysis

Adjustment variables were selected a priori based on clinical relevance and prior literature. These included age, modelled in five-year bands; sex; continent of birth as a proxy for migration background; and urbanisation category, defined according to the Italian National Institute of Statistics classification of municipalities as urban, suburban, or rural. ^28^ Comorbidity burden was assessed using the Multisource Comorbidity Score, a validated index derived from hospital discharge diagnoses and pharmaceutical prescription data.^29^ The score was categorised into predefined groups reflecting increasing morbidity burden. Extended adjustment models also incorporated variables reflecting healthcare utilisation and infection history before baseline. These included the number of SARS-CoV-2 PCR tests in the preceding six months, the number of positive tests, time since the last positive test, previous COVID-19 hospitalisation, non-COVID hospitalisation in the preceding year, and the regional health authority (ATS) of residence. We first examined determinants of undervaccination at baseline (June 1, 2022) using multivariable logistic regression models. Odds ratios and 95% confidence intervals were estimated for associations between covariates and being behind the recommended vaccination schedule. We also conducted an extended analysis using Lasso regression with a larger set of 60 covariates. ^30^ For outcome analyses, we fitted Cox proportional hazards models with time-to-event as the dependent variable. Analyses were stratified by age group (<60 years and ≥60 years), reflecting differences in recommended dose schedules and baseline risk. Vaccine deficit was included as a time-dependent categorical exposure. Individuals were censored at non-COVID death (for severe COVID-19 and long COVID analyses), deregistration, end of follow-up, or occurrence of the event. Models were first adjusted for age, sex, continent of birth, urbanisation category, and comorbidity score. Extended models incorporated additional variables on healthcare utilisation and infection history to assess robustness to potential confounding. Proportional hazards assumptions were evaluated using Schoenfeld residuals and inspection of log–log survival plots. To explore potential population-level implications of vaccination coverage, we conducted counterfactual simulations based on fitted Cox models. Under hypothetical scenarios in which all individuals were assigned to specific vaccination states (e.g., no vaccination, primary cycle only, one booster, two boosters in adults aged ≥60 years), we estimated the expected numbers of events during follow-up. These projections were compared with observed event counts. Because simulations were based on observational associations, results were interpreted cautiously as model-based projections rather than causal effects.

All analyses were performed within the secure regional computing environment using R software (version 4.3.0). Two-sided p-values below 0·05 were considered statistically significant.

## Results

As of June 1, 2022, 24·4% of the study population had not received the three recommended vaccine doses (Table 1). Overall, 9·2% were unvaccinated, 1·3% had received a single dose, and 13·9% had completed a primary course without a booster dose. Across age groups, vaccination initiation was generally followed by completion of the primary course, with few individuals remaining partially vaccinated with only one dose. The prevalence of undervaccination was greater in younger age groups, individuals born outside Europe, women, those with higher comorbidity burden, and residents of rural areas (Table 1). Temporal trends in vaccine uptake during follow-up are shown in Appendix p5. In multivariable logistic regression analyses, younger age was associated with a progressively higher probability of undervaccination, except among individuals aged 85 years or older (Figure 1; Appendix p6). Female sex, non-European country of birth, greater comorbidity burden, and rural residence were independently associated with a higher probability of undervaccination, whereas moderate comorbidity and suburban residence were associated with a lower probability. In extended models incorporating additional clinical and provider-level variables, hyperlipidaemia, renal dialysis, and older general practitioner age were associated with a lower probability of undervaccination, whereas dementia, venous thromboembolism, heart failure, and a greater number of previous COVID-19 hospitalisations were associated with a higher probability (Appendix p7-9). Residence in Bergamo, the area most severely affected during the first pandemic wave, remained independently associated with a lower probability of undervaccination.

**Figure 1:**
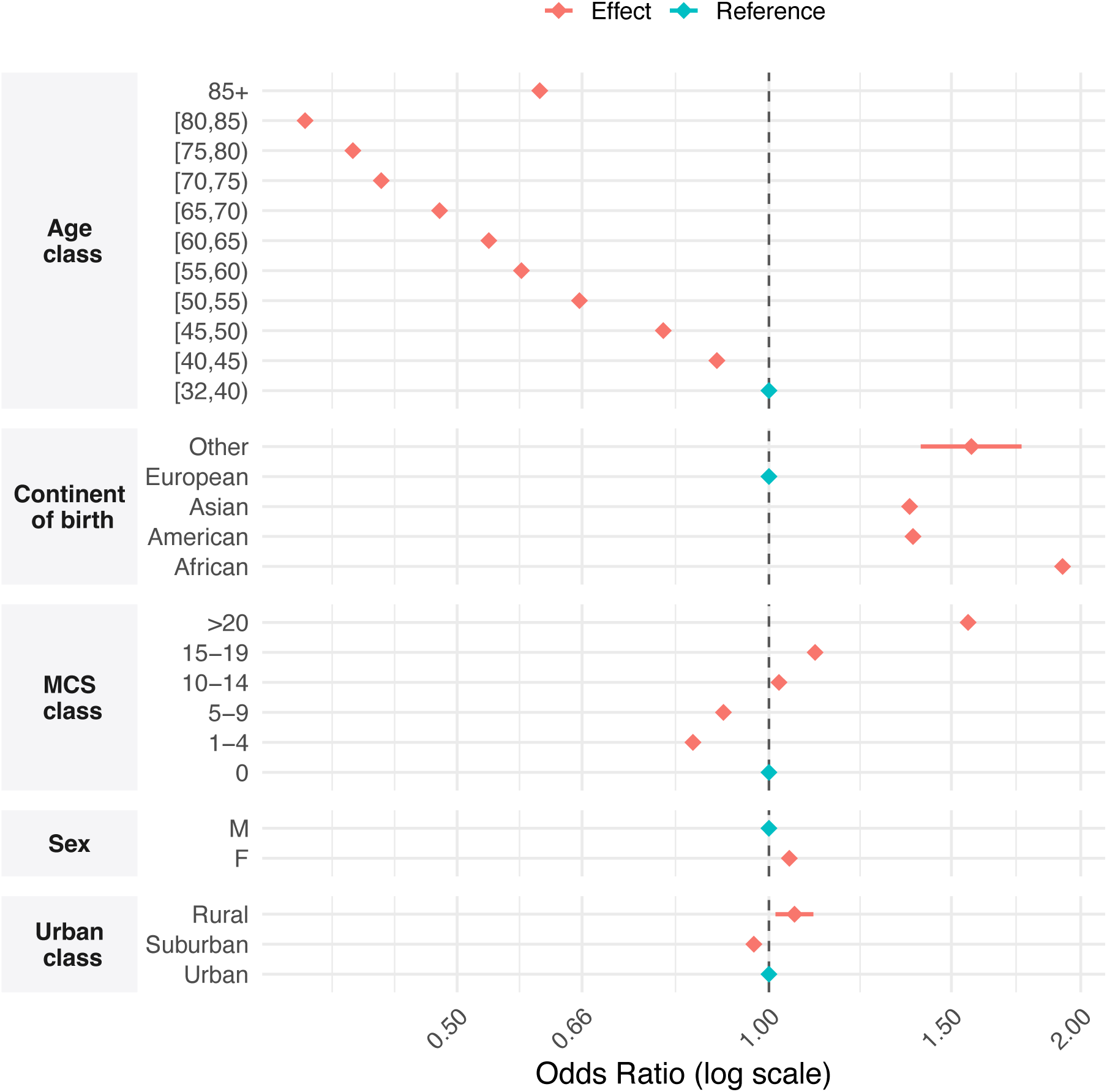
Forest plot of odds ratios of being undervaccinated at June 1 202 with 95% confidence intervals for undervaccination. Adjustments were included for age group, sex, continent of birth, urbanisation classification, and comorbidity score (MCS).

For outcome analyses, 2 899 447 individuals aged 60 years or older (2 677 004 in the long COVID analysis) and 3 842 799 individuals younger than 60 years (3 635 531 in the long COVID analysis) were included. During follow-up, 265 383 deaths were recorded (17 566 among those aged <60 years and 247 817 among those aged ≥60 years), together with 52 121 severe COVID-19 events (7 780 and 44 341, respectively) and 23 780 long COVID events (11 155 and 12 625, respectively). The incidence of recorded long COVID-related diagnoses was substantially higher in the post-pandemic period than in 2012–19 (Appendix p29). Adjusted hazard ratios (HRs) are presented in Figure 2 and Table 2. Among individuals younger than 60 years, vaccine deficit was associated with increased risk of all-cause mortality, with evidence of a dose–response gradient. Compared with those who received all recommended doses, HRs for death increased with each additional missing dose (Appendix p12 and p20-21). For severe COVID-19, a one-dose deficit was associated with increased risk (HR 1·16, 95% CI 1·11–1·21), whereas estimates for larger deficits were less precise because of small numbers of events (Appendix p13 and p21-22). In analyses of long COVID, one- and two-dose deficits were associated with increased risks (HR 1·13, 95% CI 1·07–1·19 and 1·31, 1·21–1·43, respectively), whereas the HR for complete non-vaccination did not differ significantly from that for full vaccination (Appendix p14 and p22-23).

**Figure 2:**
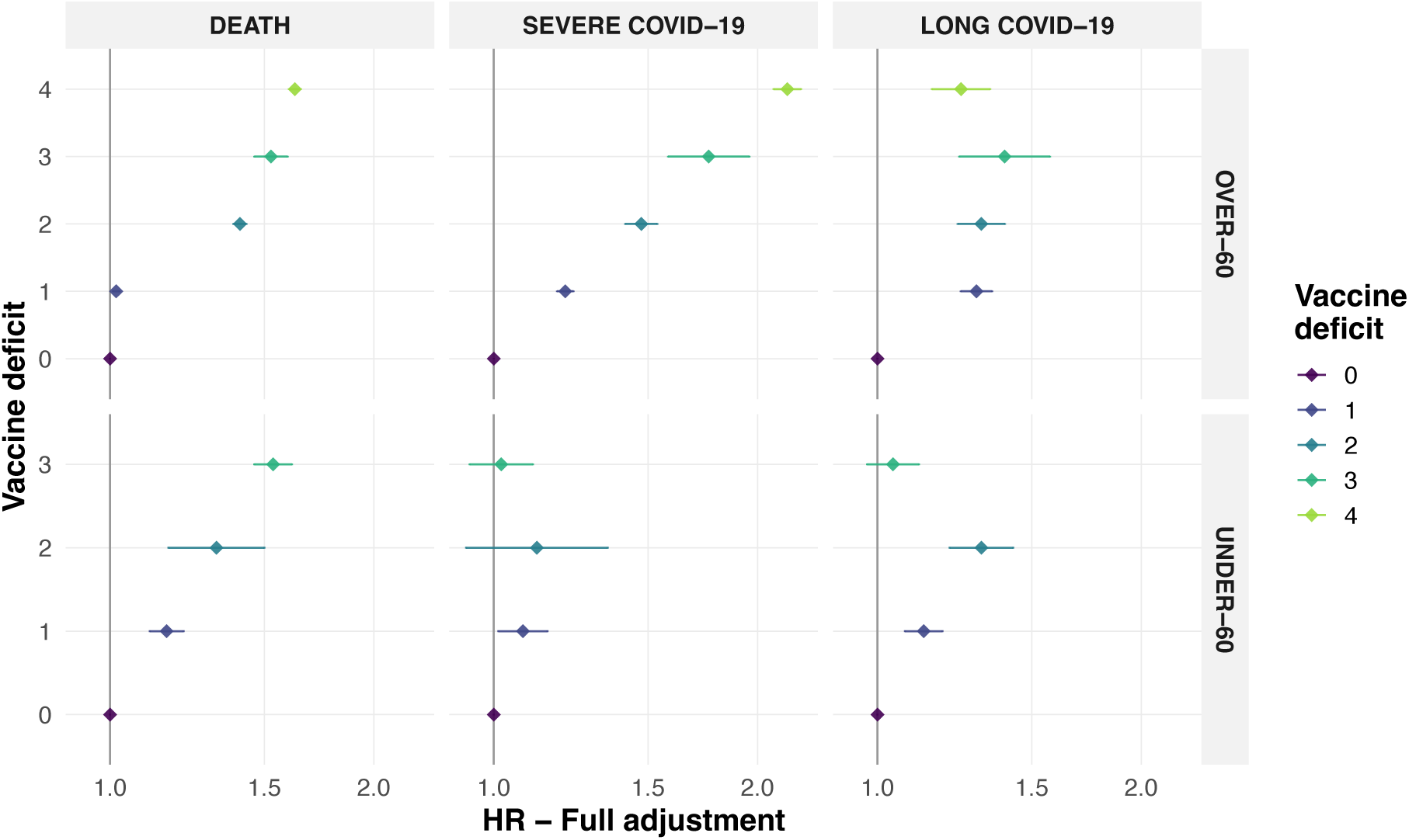
HR of the fully-adjusted analysis for mortality, severe covid-19 and long covid-19 outcome. HR = hazard ratio

**Table 2:** Events counts, follow-up person-years (per 1,000 persons), crude event rate, unadjusted HR (95% CI) and fully adjusted HR (95% CI). HR = hazard ratio.

| Vaccine deficit | Number of events | Person-time, per 1000 person-years | Event rate, per 1000 person-years | Unadjusted HR (95%CI) | Full Adjusted HR (95%CI) |
| --- | --- | --- | --- | --- | --- |
| **Under 60 - Death** |  |  |  |  |  |
| 0 | 13 032 | 7 342 | 1.78 | Reference | Reference |
| 1 | 2 416 | 1 421 | 1.70 | 0.96 (0.92-1.00) | 1.16 (1.11-1.21) |
| 2 | 257 | 128 | 2.00 | 1.13 (1.00-1.28) | 1.32 (1.17-1.50) |
| 3 | 1 861 | 915 | 2.03 | 1.15 (1.09-1.20) | 1.54 (1.46-1.61) |
| **Under 60 - Severe COVID-19** |  |  |  |  |  |
| 0 | 5 848 | 7 333 | 0.80 | Reference | Reference |
| 1 | 1 181 | 1 420 | 0.83 | 1.03 (0.96-1.09) | 1.08 (1.01-1.15) |
| 2 | 117 | 128 | 0.91 | 1.13 (0.94-1.36) | 1.12 (0.93-1.35) |
| 3 | 634 | 914 | 0.69 | 0.86 (0.79-0.93) | 1.02 (0.94-1.11) |
| **Under 60 - Long COVID-19** |  |  |  |  |  |
| 0 | 6 917 | 6 948 | 1.00 | Reference | Reference |
| 1 | 2 611 | 1 335 | 1.96 | 1.97 (1.88-2.06) | 1.13 (1.07-1.19) |
| 2 | 616 | 117 | 5.25 | 5.27 (4.85-5.72) | 1.31 (1.21-1.43) |
| 3 | 1 011 | 865 | 1.17 | 1.18 (1.10-1.26) | 1.04 (0.97-1.12) |
| **Over 60 - Death** |  |  |  |  |  |
| 0 | 112 691 | 2 536 | 44.46 | Reference | Reference |
| 1 | 99 302 | 3 640 | 27.30 | 0.59 (0.59-0.60) | 1.02 (1.01-1.03) |
| 2 | 14 931 | 457 | 32.69 | 0.72 (0.71-0.73) | 1.41 (1.38-1.43) |
| 3 | 2 054 | 49 | 42.01 | 0.92 (0.88-0.97) | 1.53 (1.46-1.60) |
| 4 | 18 839 | 448 | 42.13 | 0.93 (0.91-0.94) | 1.63 (1.60-1.65) |
| **Over 60 - Severe COVID-19** |  |  |  |  |  |
| 0 | 15 593 | 2 519 | 6.20 | Reference | Reference |
| 1 | 21 954 | 3 618 | 6.07 | 0.78 (0.76-0.80) | 1.21 (1.18-1.23) |
| 2 | 2 622 | 454 | 5.78 | 0.78 (0.75-0.82) | 1.47 (1.41-1.54) |
| 3 | 351 | 49 | 7.23 | 1.00 (0.90-1.11) | 1.76 (1.58-1.96) |
| 4 | 3 821 | 444 | 8.61 | 1.19 (1.15-1.23) | 2.16 (2.09-2.24) |
| **Over 60 - Long COVID-19** |  |  |  |  |  |
| 0 | 4 338 | 2 331 | 1.86 | Reference | Reference |
| 1 | 5 751 | 3 386 | 1.70 | 0.93 (0.89-0.97) | 1.30 (1.24-1.35) |
| 2 | 1 438 | 422 | 3.41 | 1.85 (1.75-1.97) | 1.31 (1.23-1.40) |
| 3 | 295 | 44 | 6.71 | 3.64 (3.24-4.10) | 1.40 (1.24-1.57) |
| 4 | 803 | 417 | 1.93 | 1.04 (0.97-1.13) | 1.25 (1.15-1.35) |

Among individuals aged 60 years or older, vaccine deficit was associated with increased hazards of mortality, severe COVID-19, and long COVID (Figure 2). For mortality and severe COVID-19, associations demonstrated a clear gradient, with progressively higher HRs as the number of missing doses increased (Appendix p16 and p25-26). In severe COVID-19 analyses, HRs ranged from 1·21 (95% CI 1·18–1·23) for a one-dose deficit to 2·16 (2·09–2·24) for a four-dose deficit (Appendix p17 and p26-27). For long COVID, all vaccine-deficit categories were associated with moderately increased risks, with less evidence of a marked dose–response pattern (Appendix p18 and p27-28). Age and comorbidity were independently associated with all three outcomes. Among individuals younger than 60 years, increasing age was associated with higher risks of mortality and severe COVID-19 but lower hazard of long COVID. In those aged 60 years or older, age was associated with mortality, severe COVID-19, and long COVID. Higher comorbidity scores were consistently associated with increased risks across subgroups. Female sex was associated with lower risks of mortality and severe COVID-19 but not with reduced hazard of long COVID in the younger subgroup. The basic adjustment analysis shows little to no difference between the estimates for mortality and severe COVID-19 outcomes between basic and additional adjustment except for the long COVID-19 analysis (Appendix p30), confirming the sensibility of the latter outcome found in the sensitivity analysis (Appendix p31).

In model-based projections counterfactual analyses (Appendix p32–33), a hypothetical scenario of complete non-vaccination among individuals younger than 60 years was associated with an estimated 7 083 excess deaths compared with observed uptake, whereas universal booster coverage was estimated to prevent 1 892 fewer long COVID cases. Among those aged 60 years or older, a primary-course-only scenario estimated 68 404 excess deaths and complete non-vaccination 96 276 excess deaths. By contrast, universal first- or second-booster coverage was estimated to prevent 10 971 and 11 711 fewer deaths, respectively. For severe COVID-19 in older individuals, complete non-vaccination was estimated to result in 39 547 excess cases, whereas universal second-booster coverage was estimated to result in 2 964 fewer cases. For long COVID, the primary-course-only scenario estimated 17 506 excess cases, whereas universal second-booster coverage estimated 1 457 fewer cases.

## Discussion

In this population-based study of 6·8 million adults residing in Lombardy, we characterised patterns of COVID-19 undervaccination and quantified associations between vaccine-dose deficit and all-cause mortality, severe COVID-19, and post-COVID conditions. Undervaccination was common: by June 1, 2022, almost one in four adults had not received the recommended three doses. Undervaccination was higher in specific population groups, including younger adults, individuals born outside Europe, residents of rural areas, and those with higher comorbidity burden. Vaccine-dose deficit was associated with higher risks of adverse outcomes, with dose–response gradients for mortality and severe COVID-19 among adults aged 60 years or older. For long COVID, associations in older adults were consistent with a protective effect of complete primary and booster vaccination.

The sociodemographic patterning of undervaccination in our study is consistent with findings from other large register-based analyses in northern Europe and the UK, which have highlighted the role of migration background, socioeconomic position, and risk perception in shaping vaccine uptake. ^8–10^ The persistence of these gradients across health systems suggests structural barriers, such as differential access to information, language, trust, and service organisation, rather than purely individual choice. ^11^ The lower prevalence of undervaccination observed in the Bergamo area, and among individuals with previous COVID-19 hospitalisation, plausibly reflects the salience of first-hand pandemic experience during the initial wave, which may have increased perceived risk and motivation to vaccinate. Associations between certain cardiovascular conditions and undervaccination might reflect contemporaneous media coverage and public concern about vaccine-related cardiovascular events, although residual confounding cannot be excluded. ^31,32^

Associations between vaccine-dose deficit and risks of severe COVID-19 and death, particularly among adults aged 60 years or older, are consistent with evidence from meta-analyses and national cohort studies showing that incomplete vaccination substantially increases the risk of hospitalisation and mortality. ^9,17^ Our findings extend this evidence to an Italian population, with time-varying exposure modelling and follow-up spanning multiple variant waves into late 2024. The magnitude of association was greatest among older adults and those with multimorbidity, consistent with biological plausibility and the well-established age gradient in COVID-19 severity. In counterfactual analyses, we estimated substantial excess deaths and severe cases under scenarios of incomplete or absent vaccination, and additional gains under universal booster coverage.^7^ Although such simulations rely on model assumptions, they provide an interpretable measure of the potential population impact of vaccination programmes in a real-world regional context.

Associations with long COVID were more heterogeneous. Among adults aged 60 years or older, increasing vaccine-dose deficits were associated with a higher risk of recorded long-COVID symptoms, supporting a protective effect of complete primary and booster vaccination schedules. Among younger adults, effect estimates were smaller and not consistent. These findings underscore the susceptibility of long-COVID endpoints derived from routine care to changes in testing availability, healthcare–seeking behaviour, and coding practices over time.^27^ They are nonetheless broadly consistent with a heterogeneous literature in which most large studies report that vaccination before infection reduces the risk of post-COVID condition, albeit with effect sizes that vary by variant period, follow-up duration, and outcome definition. ^21,23^ Our symptom-based definition was anchored to the WHO case definition, but routine data inevitably capture only a subset of patients who present for care and receive a coded diagnosis.

Strengths of this study include near-complete coverage of a large European region; individual-level linkage across vaccination, testing, hospital, primary and community care, and mortality registers; extended follow-up across successive variant periods; and modelling of vaccination status as a time-varying exposure with comprehensive adjustment for demographic, clinical, and healthcare-use characteristics. Limitations are inherent to observational analyses of administrative data. Residual confounding and healthy-vaccinee effects may persist despite adjustment. ^33^ Our definitions of severe COVID-19 and mortality required pragmatic operational choices, including inclusion of deaths within two months of a positive test to mitigate incomplete cause-of-death information. Ascertainment of long COVID relied on healthcare contacts and symptom coding, and changing testing policies during later phases of the pandemic likely introduced selection bias and misclassification, as reflected in the sensitivity analyses. Finally, some highly frail individuals may fall into vaccination recommendation categories different from those captured for the overall population, particularly among those under 60; however, they are likely to represent only a very small proportion of the population.

## Conclusions

Taken together, and consistent with existing evidence, our findings reinforce the public health importance of sustaining high, up-to-date vaccination coverage to prevent severe COVID-19 and death and to mitigate longer-term sequelae. Maintaining timely booster administration, particularly among older adults and individuals with multimorbidity, should remain central to vaccination strategies, with programmes designed to anticipate waning immunity and extend proactive outreach beyond completion of the primary series. At the same time, reducing persistent undervaccination will require targeted approaches for groups with lower uptake, including younger adults, migrants, and residents of rural communities, with interventions that reduce structural barriers and strengthen engagement with health services.

## List of abbreviations

CI: Confidence interval
COVID-19: Coronavirus disease 2019
HR: Hazard ratio
ICD-10: International Classification of Diseases, 10th Revision
MCS: Multisource Comorbidity Score
PCR: Polymerase chain reaction
SARS-CoV-2: Severe acute respiratory syndrome coronavirus 2
WHO: World Health Organization

## Declarations

### Ethics approval and consent to participate

Not applicable

### Consent for publication

Not applicable

## Authors’ contributions

AC, EDA and FI conceived this study. AC and KML accessed and verified the data. AC drafted the statistical analysis plan and led the analysis. AC and KML accessed and verified the data. AC, FI, KML, and EDA drafted and revised the manuscript. All authors review and approved the manuscript before submission. EDA and FI are Co-Heads of the Health Data Science Centre of Human Technopole and coordinated approvals for and access to data within the Lombardy Region ARIA service for the COV-CVD project. EDA was responsible for the final decision to submit the manuscript.

## Competing interests

All authors declare no competing interests.

## Data Availability

Data availability for Lombardy region data is publicly available on the Epidemiological Observatory of the Lombardy Region website (www.osservatorioepidemiologico.regione.lombardia.it/wps/portal/site/osservatorio-epidemiologico/DettaglioRedazionale/collaborazioni-con-gli-enti/daas+2-0/red-daas-2-0). Access to the Lombardy region data can be obtained by submitting a project application for individual-level data. The application includes information on the purpose of data use; the requested data, including variables and definitions of the target and control groups; the required data dates; and a data utilisation plan. The requests are evaluated on a case-by-case basis. Once approved, the data are sent to a secure computing environment (Daas 2.0). The analysis code used to produce the results is available on GitHub at: https://github.com/ht-diva/UndervaccinationRL.git.

## Acknowledgments

We are deeply grateful to all individuals in the Lombardy Region, whose data made this study possible. We would also like to acknowledge the entire Aria team and Osservatorio Epidemiologico Lombardo for making the data available for the study. The present research is part of the activities of “Dipartimento di Eccellenza 2023-2027”. We would also like to acknowledge Claudia Giambartolemi for her suggestions on the draft of the manuscript.

## Funding

No fundings to disclose

## Notes

### Competing Interest Statement

The authors have declared no competing interest.

### Author Declarations

Data availability for Lombardy region data is publicly available on the Epidemiological Observatory of the Lombardy Region website (www.osservatorioepidemiologico.regione.lombardia.it/wps/portal/site/osservatorio-epidemiologico/DettaglioRedazionale/collaborazioni-con-gli-enti/daas+2-0/red-daas-2-0). Access to the Lombardy region data can be obtained by submitting a project application for individual-level data. The application includes information on the purpose of data use; the requested data, including variables and definitions of the target and control groups; the required data dates; and a data utilisation plan. The requests are evaluated on a case-by-case basis. Once approved, the data are sent to a secure computing environment (Daas 2.0). The datasets used in our study were individual-level data and they had been de-identified prior to use.

