## Appendix for "Determinants and long-term outcomes of COVID-19 undervaccination: a cohort study of 6.8 million individuals in Lombardy, Italy"

### Reporting checklist (STROBE)

|  | Item No | Recommendation | Page No |
| --- | --- | --- | --- |
| **Title and abstract** | 1 | (*a*) Indicate the study’s design with a commonly used term in the title or the abstract | 1 |
|  |  | (*b*) Provide in the abstract an informative and balanced summary of what was done and what was found | 1-2 |
| Introduction | | | |
| Background/rationale | 2 | Explain the scientific background and rationale for the investigation being reported | 3-4 |
| Objectives | 3 | State-specific objectives, including any prespecified hypotheses | 3-4 |
| Methods | | | |
| Study design | 4 | Present key elements of study design early in the paper | 3-4 |
| Setting | 5 | Describe the setting, locations, and relevant dates, including periods of recruitment, exposure, follow-up, and data collection | 5 |
| Participants | 6 | (*a*) Give the eligibility criteria, and the sources and methods of selection of participants. Describe methods of follow-up | 5-6 |
|  |  | (*b*) For matched studies, give matching criteria and number of exposed and unexposed |  |
| Variables | 7 | Clearly define all outcomes, exposures, predictors, potential confounders, and effect modifiers. Give diagnostic criteria, if applicable | 6-7 |
| Data sources/ measurement | 8* | For each variable of interest, give sources of data and details of methods of assessment (measurement). Describe comparability of assessment methods if there is more than one group | 6-8 |
| Bias | 9 | Describe any efforts to address potential sources of bias | 8 |
| Study size | 10 | Explain how the study size was arrived at | 9-10 |
| Quantitative variables | 11 | Explain how quantitative variables were handled in the analyses. If applicable, describe which groupings were chosen and why | 6-8 |
| Statistical  methods | 12 | (*a*) Describe all statistical methods, including those used to control for confounding | 7-8 |
|  |  | (*b*) Describe any methods used to examine subgroups and interactions |  |
|  |  | (*c*) Explain how missing data were addressed | 6-8 |
|  |  | (*d*) If applicable, explain how loss to follow-up was addressed | 6-8 |
|  |  | (*e*) Describe any sensitivity analyses | 6-8 |
| Results | | |  |
| Participants | 13* | (a) Report numbers of individuals at each stage of study—eg numbers potentially eligible, examined for eligibility, confirmed eligible, included in the study, completing follow-up, and analysed | 9-10 |
|  |  | (b) Give reasons for non-participation at each stage |  |
|  |  | (c) Consider the use of a flow diagram |  |
| Descriptive data | 14* | (a) Give characteristics of study participants (eg demographic, clinical, social) and information on exposures and potential confounders | 9-11 |
|  |  | (b) Indicate the number of participants with missing data for each variable of interest | 9-11 |
|  |  | (c) Summarise follow-up time (eg, average and total amount) |  |
| Outcome data | 15* | Report numbers of outcome events or summary measures over time | 9-12 |
| Main results | 16 | (*a*) Give unadjusted estimates and, if applicable, confounder-adjusted estimates and their precision (eg, 95% confidence interval). Make clear which confounders were adjusted for and why they were included | 9-12 |
|  |  | (*b*) Report category boundaries when continuous variables were categorised |  |
|  |  | (*c*) If relevant, consider translating estimates of relative risk into absolute risk for a meaningful time period |  |
| Other analyses | 17 | Report other analyses done—eg analyses of subgroups and interactions, and sensitivity analyses | 9-12 |
| Discussion |  |  |  |
| Key results | 18 | Summarise key results with reference to study objectives | 12-13 |
| Limitations | 19 | Discuss limitations of the study, taking into account sources of potential bias or imprecision. Discuss both direction and magnitude of any potential bias | 13-14 |
| Interpretation | 20 | Give a cautious overall interpretation of results considering objectives, limitations, multiplicity of analyses, results from similar studies, and other relevant evidence | 13-14 |
| Generalisability | 21 | Discuss the generalisability (external validity) of the study results | 15-16 |
| Other information |  |  |  |
| Funding | 22 | Give the source of funding and the role of the funders for the present study and, if applicable, for the original study on which the present article is based | 18 |

Table S1: Reporting checklist (STROBE) for observational studies

### Vaccination uptake / coverage

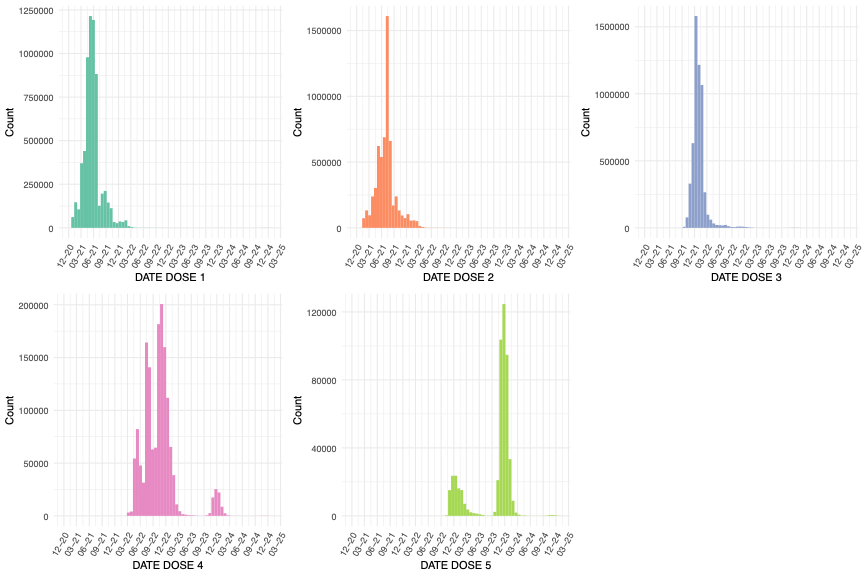

Figure S1: Distribution of dose delivery by dose number from December 2020 to December 2024

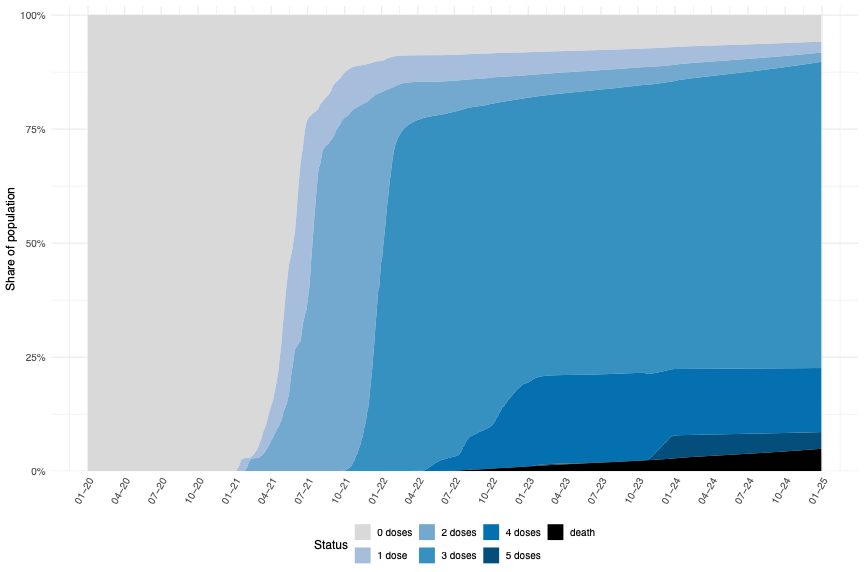

Figure S2: Vaccination status of the population from 2020-1-1 to 2024-12-31 by doses received

### Determinants of undervaccination

#### Undervaccination multivariate logistic regression analysis

| **Variable** | **Adjusted OR (95% CI)** |
| --- | --- |
| **MCS class** | |
| 0 | Reference |
| 1-4 | 0·84 (0·84-0·85) |
| 5-9 | 0·90 (0·90-0·91) |
| 10-14 | 1·02 (1·01-1·03) |
| 15-19 | 1·11 (1·09-1·12) |
| >20 | 1·56 (1·54-1·58) |
| **Continent of birth** | |
| Europe | Reference |
| Africa | 1·92 (1·90-1·94) |
| American | 1·38 (1·36-1·39) |
| Asian | 1·37 (1·35-1·38) |
| Other | 1·57 (1·40-1·75) |
| **Sex** | |
| Male | Reference |
| Female | 1·05 (1·04-1·05) |
| **Urbanisation class** | |
| Urban | Reference |
| Suburban | 0·97 (0·96-0·97) |
| Rural | 1·06 (1·01-1·10) |
| **Age class** | |
| [32,40) | Reference |
| [40,45) | 0·89 (0·88-0·90) |
| [45,50) | 0·79 (0·79-0·80) |
| [50,55) | 0·66 (0·65-0·66) |
| [55,60) | 0·58 (0·57-0·58) |
| [60,65) | 0·54 (0·53-0·54) |
| [65,70) | 0·48 (0·48-0·48) |
| [70,75) | 0·42 (0·42-0·43) |
| [75,80) | 0·40 (0·39-0·40) |
| [80,85) | 0·36 (0·35-0·36) |
| [85, 105) | 0·60 (0·60-0·61) |

Table S2: Undervaccination OR and 95% CI. OR = odds ratio

#### Undervaccination Lasso logistic regression analysis

Lasso results are from a penalised logistic regression fitted to baseline undervaccination, with the penalty selected by 9-fold cross-validation (penalty = 0·00115139539932645; cross-validated AUC = 0·627). Coefficients (direction + scale).

Coefficients are on the log-odds scale: positive values indicate higher odds of undervaccination and negative values indicate lower odds (e.g., “Age 32–40” has a positive coefficient, whereas “Continent of Birth: Europe” is negative Table S3 and S4). Here, Lasso is presented to rank a larger candidate set of determinants (Table S5 shows coefficients shrunk to zero) and is not intended for inferential interpretation.

Predictors are reported as indicator variables using the labels shown in the tables (e.g., “Age 32–40”, “Last PCR Test = 3–4 weeks”, “Continent of Birth: Europe”, “ATS Bergamo”). Each coefficient, therefore, compares the presence vs absence of that labelled feature (implicit reference is everyone not meeting that label).

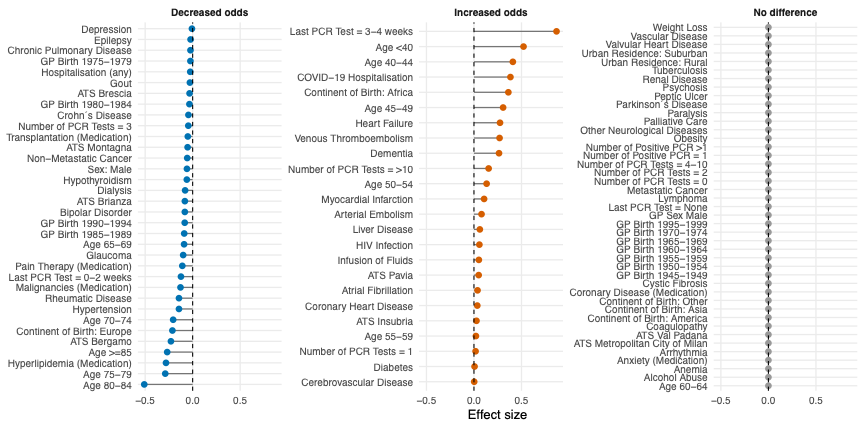

Figure S3: Lasso coefficients lollipop plot

| **Lasso coefficient** | **Prevalence** | **Covariate** |
| --- | --- | --- |
| -0·50913 | 6·17% | Age 80–84 |
| -0·28834 | 7·13% | Age 75–79 |
| -0·28052 | 17·88% | Hyperlipidemia (Medication) |
| -0·26794 | 5·98% | Age ≥85 |
| -0·22857 | 10·60% | ATS Bergamo |
| -0·21312 | 93·17% | Continent of Birth: Europe |
| -0·20516 | 8·06% | Age 70–74 |
| -0·14404 | 29·75% | Hypertension |
| -0·144 | 0·92% | Rheumatic Disease |
| -0·12794 | 2·40% | Malignancies (Medication) |
| -0·12311 | 1·00% | Last PCR Test = 0-2 weeks |
| -0·10873 | 29·50% | Pain Therapy (Medication) |
| -0·09978 | 2·69% | Glaucoma |
| -0·09022 | 8·51% | Age 65–69 |
| -0·08886 | 5·80% | GP Birth 1985–1989 |
| -0·08354 | 1·76% | GP Birth 1990–1994 |
| -0·08232 | 0·20% | Bipolar Disorder |
| -0·082 | 11·77% | ATS Brianza |
| -0·07933 | 0·03% | Dialysis |
| -0·06135 | 5·42% | Hypothyroidism |
| -0·06001 | 47·39% | Sex: Male |
| -0·05715 | 0·77% | Non-Metastatic Cancer |
| -0·05319 | 3·02% | ATS Montagna |
| -0·05096 | 0·25% | Transplantation (Medication) |
| -0·0457 | 0·54% | Number of PCR Tests = 3 |
| -0·04447 | 2·08% | Crohn’s Disease |
| -0·03399 | 5·47% | GP Birth 1980–1984 |
| -0·03249 | 11·71% | ATS Brescia |
| -0·02895 | 3·65% | Gout |
| -0·02529 | 24·47% | Hospitalisation (any) |
| -0·02364 | 3·44% | GP Birth 1975–1979 |
| -0·02306 | 14·59% | Chronic Pulmonary Disease |
| -0·0219 | 4·02% | Epilepsy |
| -0·00753 | 1·72% | Depression |

Table S3: Lasso undervaccination negative coefficients and their population prevalence (non-zero)

| **Lasso coefficient** | **Prevalence** | **Covariate** |
| --- | --- | --- |
| 0·868478 | 0·71% | Last PCR Test = 3-4 weeks |
| 0·521405 | 9·85% | Age <40 |
| 0·409056 | 8·78% | Age 40–44 |
| 0·383651 | 0·64% | COVID-19 Hospitalisation |
| 0·36176 | 2·74% | Continent of Birth: Africa |
| 0·306555 | 11·15% | Age 45–49 |
| 0·273967 | 1·73% | Heart Failure |
| 0·269275 | 0·64% | Venous Thromboembolism |
| 0·263474 | 0·10% | Dementia |
| 0·154338 | 0·07% | Number of PCR Tests = >10 |
| 0·133097 | 12·03% | Age 50–54 |
| 0·106644 | 1·60% | Myocardial Infarction |
| 0·079618 | 0·16% | Arterial Embolism |
| 0·061791 | 3·14% | Liver Disease |
| 0·056942 | 0·23% | HIV Infection |
| 0·051366 | 0·26% | Infusion of Fluids |
| 0·049523 | 5·61% | ATS Pavia |
| 0·037612 | 2·33% | Atrial Fibrillation |
| 0·035023 | 3·97% | Coronary Heart Disease |
| 0·026457 | 14·81% | ATS Insubria |
| 0·020457 | 12·14% | Age 55–59 |
| 0·016973 | 4·95% | Number of PCR Tests = 1 |
| 0·006219 | 8·83% | Diabetes |
| 0·002496 | 0·46% | Cerebrovascular Disease |

Table S4: Lasso undervaccination positive coefficients and their population prevalence (non-zero)

| **Lasso coefficient** | **Prevalence** | **Covariate** |
| --- | --- | --- |
| 0 | 98·30% | Last PCR Test = None |
| 0 | 92·66% | Number of PCR Tests = 0 |
| 0 | 57·30% | GP Sex Male |
| 0 | 44·45% | Urban Residence: Suburban |
| 0 | 35·35% | ATS Metropolitan City of Milan |
| 0 | 32·75% | GP Birth 1955–1959 |
| 0 | 24·37% | GP Birth 1960–1964 |
| 0 | 15·64% | Coronary Disease (Medication) |
| 0 | 12·57% | GP Birth 1950–1954 |
| 0 | 10·21% | Age 60–64 |
| 0 | 9·96% | GP Birth 1965–1969 |
| 0 | 9·78% | Peptic Ulcer |
| 0 | 7·15% | ATS Val Padana |
| 0 | 5·38% | Arrhythmia |
| 0 | 3·90% | Valvular Heart Disease |
| 0 | 3·88% | GP Birth 1970–1974 |
| 0 | 2·51% | Vascular Disease |
| 0 | 2·27% | Continent of Birth: Asia |
| 0 | 1·80% | Continent of Birth: America |
| 0 | 1·21% | Number of PCR Tests = 2 |
| 0 | 1·21% | Number of Positive PCR = 1 |
| 0 | 1·07% | Psychosis |
| 0 | 1·03% | Anemia |
| 0 | 0·77% | Parkinson’s Disease |
| 0 | 0·57% | Number of PCR Tests = 4-10 |
| 0 | 0·50% | Number of Positive PCR >1 |
| 0 | 0·46% | Palliative Care |
| 0 | 0·42% | Renal Disease |
| 0 | 0·30% | Other Neurological Diseases |
| 0 | 0·29% | Obesity |
| 0 | 0·28% | Metastatic Cancer |
| 0 | 0·27% | Cystic Fibrosis |
| 0 | 0·16% | Paralysis |
| 0 | 0·12% | Lymphoma |
| 0 | 0·10% | Tuberculosis |
| 0 | 0·09% | Urban Residence: Rural |
| 0 | 0·07% | Alcohol Abuse |
| 0 | 0·05% | Coagulopathy |
| 0 | 0·04% | Anxiety (Medication) |
| 0 | 0·02% | Weight Loss |
| 0 | 0·02% | Continent of Birth: Other |
| 0 | 0·00% | GP Birth 1995–1999 |
| 0 | 0·00% | GP Birth 1945–1949 |

Table S5: Lasso undervaccination coefficients and their population prevalence (shrunkinked)

### Survival analyses

This section reports time-to-event (survival) analyses assessing the association between time-varying vaccine dose deficit and subsequent risk of all-cause mortality, severe COVID-19, and long COVID symptom diagnoses during follow-up from June 1, 2022, to Dec 31, 2024.

Results are presented as hazard ratios (HRs) with 95% confidence intervals, comparing each deficit category with the “up-to-date” reference group, and we show both a basic adjustment set and a more fully adjusted set (including healthcare utilisation and infection-history variables) to evaluate robustness to potential confounding.

| **Covariate** | **Abbreviation** | **Definition** | **Coding / levels** | **Source/notes** |
| --- | --- | --- | --- | --- |
| **Basic adjustment** | | | | |
| Age | AGE CLASS | Age at baseline | **5-year bands** | Age-stratified analyses: **<60 vs ≥60** |
| Sex | SEX | Demographic sex | Female / Male |  |
| Continent of birth | CONTINENT | Proxy for migration background | Europe (ref), Africa, America, Asia, Other |  |
| Urbanisation category | URBAN DOMICILE | Municipality classification (ISTAT) | Urban (ref), Suburban, Rural |  |
| Multisource Comorbidity Score (MCS) | MCS CLASS | Validated comorbidity index from hospital discharge diagnoses and pharmaceutical prescriptions | Categorised:   - **0 (0)** - **1–4 (1)** - **5–9 (2)** - **10–14 (3)** - **15–19 (4)** - **>20 (5)** |  |
| **Full adjustment** | | | | |
| ATS of residence | ATS | Regional health authority (ATS) of residence | - DELL'INSUBRIA - DELLA BRIANZA - DI MILANO (ref) - DELLA MONTAGNA - DELLA VAL PADANA - DI BERGAMO, - DI BRESCIA, - DI PAVIA |  |
| Number of PCR tests | N PCR | Count of SARS-CoV-2 PCR tests in the **preceding 6 months** | Categorised:   - **0 (0),** - **1 (1)** - **2 (2)** - **3 (3),** - **4–9 (4)** - **≥10 (5)** |  |
| Number of positive PCR tests | N PCR POS | Count of positive PCR tests in the **preceding 6 months** | Categorised :   - **0 (0)** - **1 (1)** - **≥2 (2)** |  |
| Weeks to last positive PCR | WEEKS TO LAST POSITIVE PCR | Time since the last positive SARS-CoV-2 PCR test | Categorised:   - **None (0)** - **0–13 (1)** - **14–26 (2)** - **≥27** (3) |  |
| Previous COVID-19 hospitalisation | C-HOSP | Prior COVID-19 hospitalisation before baseline | 0 vs ≥1 |  |
| Non-COVID hospitalisation | HOSP | Non-COVID hospitalisation in the **preceding year** | 0 vs ≥1 |  |

Table S6: Covariates definition in basic and full adjustment survival analysis

#### Basic adjustment analysis

##### Under 60 - Basic adjustment coefficients plot

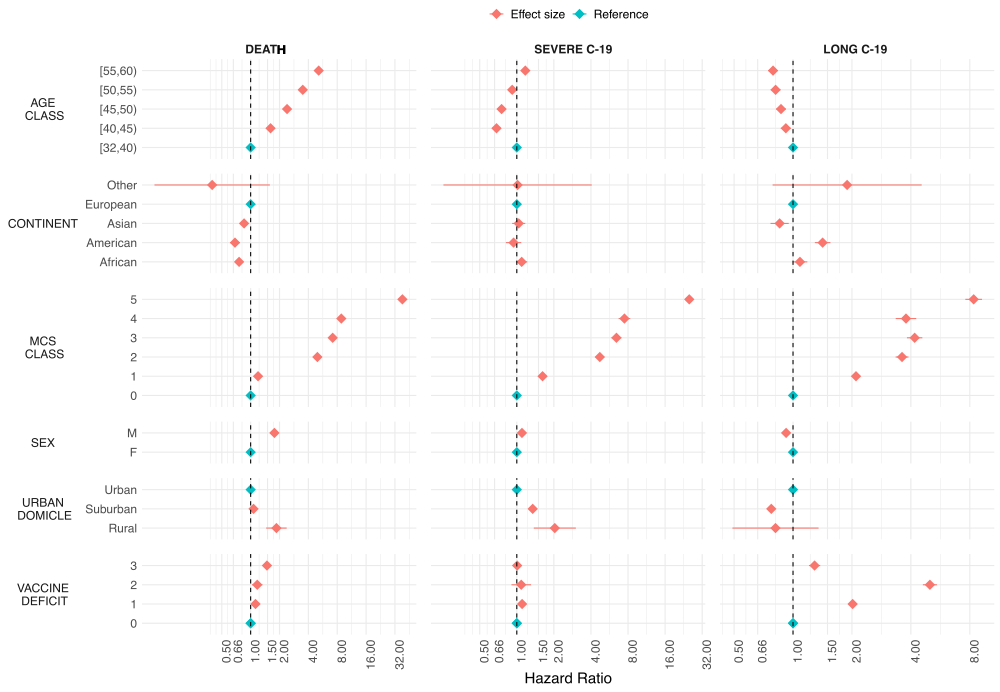

Figure S4: Forest plot basic adjustment analysis – under-60

##### Under 60 - Death outcome coefficients

| **Variable** | **Level** | **Events** | **Person time** | **Annual event rate**  **x1000 individuals** | **Adjusted HR**  **(95% CI)** |
| --- | --- | --- | --- | --- | --- |
| Age class | [32,40) | 1079 | 768078888 | 0·51 | Ref |
| Age class | [40,45) | 1388 | 588985796 | 0·86 | 1·61 (1·49-1·75) |
| Age class | [45,50) | 2767 | 726340935 | 1·39 | 2·40 (2·23-2·57) |
| Age class | [50,55) | 4699 | 758117494 | 2·26 | 3·50 (3·27-3·74) |
| Age class | [55,60) | 7633 | 738011487 | 3·78 | 5·12 (4·81-5·47) |
| Continent | African | 572 | 154974405 | 1·35 | 0·76 (0·70-0·82) |
| Continent | American | 282 | 102552859 | 1·00 | 0·69 (0·61-0·77) |
| Continent | Asian | 458 | 132665727 | 1·26 | 0·85 (0·78-0·94) |
| Continent | European | 16252 | 3188449823 | 1·86 | Ref |
| Continent | Other | 2 | 891786 | 0·82 | 0·40 (0·10-1·59) |
| Vaccine deficit | 0 | 13032 | 2679698026 | 1·78 | Ref |
| Vaccine deficit | 1 | 2416 | 518812580 | 1·70 | 1·12 (1·08-1·17) |
| Vaccine deficit | 2 | 257 | 46879674 | 2·00 | 1·17 (1·04-1·33) |
| Vaccine deficit | 3 | 1861 | 334144320 | 2·03 | 1·48 (1·41-1·55) |
| MCS class | 0 | 6174 | 2300923528 | 0·98 | Ref |
| MCS class | 1-4 | 3333 | 1024309240 | 1·19 | 1·19 (1·15-1·25) |
| MCS class | 5-9 | 1813 | 119384803 | 5·54 | 4·97 (4·71-5·24) |
| MCS class | 10-14 | 1323 | 64532218 | 7·48 | 7·17 (6·76-7·61) |
| MCS class | 15-19 | 909 | 38394406 | 8·64 | 8·82 (8·23-9·46) |
| MCS class | >20 | 4014 | 31990405 | 45·80 | 38·28 (36·76-39·86) |
| Sex | F | 6581 | 1793029202 | 1·34 | Ref |
| Sex | M | 10985 | 1786505398 | 2·24 | 1·77 (1·72-1·82) |
| Urbanisation class | Rural | 64 | 4982186 | 4·69 | 1·86 (1·45-2·37) |
| Urbanisation class | Suburban | 8301 | 1621776583 | 1·87 | 1·07 (1·04-1·10) |
| Urbanisation class | Urban | 9201 | 1952775831 | 1·72 | Ref |

Table S7: Basic adjustment HR - under-60 mortality analysis. Number of events, person-time (days), event rate per 1,000 individuals per year, adjusted HR with 95% confidence intervals.

##### Under 60 - Severe COVID-19 outcome coefficients

| **Variable** | **Level** | **Events** | **Person time** | **Annual event rate**  **x1000 individuals** | **Adjusted HR**  **(95% CI)** |
| --- | --- | --- | --- | --- | --- |
| Age class | [32,40) | 1601 | 767052825 | 0·76 | Ref |
| Age class | [40,45) | 866 | 588465441 | 0·54 | 0·68 (0·63-0·74) |
| Age class | [45,50) | 1238 | 725647911 | 0·62 | 0·75 (0·70-0·81) |
| Age class | [50,55) | 1716 | 757185199 | 0·83 | 0·92 (0·86-0·98) |
| Age class | [55,60) | 2359 | 736811968 | 1·17 | 1·17 (1·10-1·25) |
| Continent of birth | African | 371 | 154760911 | 0·87 | 1·10 (0·99-1·22) |
| Continent of birth | American | 191 | 102439479 | 0·68 | 0·94 (0·81-1·09) |
| Continent of birth | Asian | 266 | 132511873 | 0·73 | 1·04 (0·92-1·17) |
| Continent of birth | European | 6950 | 3184560581 | 0·80 | Ref |
| Continent of birth | Other | 2 | 890500 | 0·82 | 1·01 (0·25-4·06) |
| Vaccine deficit | 0 | 5848 | 2676437271 | 0·80 | Ref |
| Vaccine deficit | 1 | 1181 | 518137850 | 0·83 | 1·10 (1·04-1·18) |
| Vaccine deficit | 2 | 117 | 46813526 | 0·91 | 1·09 (0·90-1·31) |
| Vaccine deficit | 3 | 634 | 333774697 | 0·69 | 1·01 (0·93-1·09) |
| MCS class | 0 | 2928 | 2299190934 | 0·46 | Ref |
| MCS class | 1-4 | 2122 | 1023076441 | 0·76 | 1·62 (1·53-1·71) |
| MCS class | 5-9 | 741 | 118986381 | 2·27 | 4·72 (4·35-5·12) |
| MCS class | 10-14 | 544 | 64238940 | 3·09 | 6·44 (5·88-7·06) |
| MCS class | 15-19 | 367 | 38195487 | 3·51 | 7·48 (6·71-8·34) |
| MCS class | >20 | 1078 | 31475161 | 12·50 | 25·14 (23·42-26·99) |
| Sex | F | 3873 | 1790763124 | 0·79 | Ref |
| Sex | M | 3907 | 1784400220 | 0·80 | 1·10 (1·05-1·15) |
| Urbanisation class | Rural | 25 | 4966531 | 1·84 | 2·04 (1·37-3·02) |
| Urbanisation class | Suburban | 4095 | 1619472915 | 0·92 | 1·35 (1·29-1·41) |
| Urbanisation class | Urban | 3660 | 1950723898 | 0·68 | Ref |

Table S8: Basic adjustment HR - under-60 severe COVID-19 analysis. Number of events, person-time (days), event rate per 1,000 individuals per year, adjusted HR with 95% confidence intervals.

##### Under 60 - Severe long COVID-19 outcome coefficients

| **Variable** | **Level** | **Events** | **Person time** | **Annual event rate**  **x1000 individuals** | **Adjusted HR**  **(95% CI)** |
| --- | --- | --- | --- | --- | --- |
| Age class | [32,40) | 2645 | 722675028 | 1·34 | Ref |
| Age class | [40,45) | 1852 | 554830228 | 1·22 | 0·92 (0·87-0·98) |
| Age class | [45,50) | 2162 | 686232467 | 1·15 | 0·87 (0·82-0·92) |
| Age class | [50,55) | 2262 | 718089980 | 1·15 | 0·81 (0·77-0·86) |
| Age class | [55,60) | 2234 | 700010970 | 1·16 | 0·79 (0·75-0·84) |
| Continent of birth | African | 542 | 141306660 | 1·40 | 1·09 (0·99-1·18) |
| Continent of birth | American | 485 | 96326032 | 1·84 | 1·42 (1·29-1·55) |
| Continent of birth | Asian | 347 | 125760251 | 1·01 | 0·85 (0·77-0·95) |
| Continent of birth | European | 9776 | 3017600368 | 1·18 | Ref |
| Continent of birth | Other | 5 | 845362 | 2·16 | 1·89 (0·79-4·54) |
| Vaccine deficit | 0 | 6917 | 2536019019 | 1·00 | Ref |
| Vaccine deficit | 1 | 2611 | 487340746 | 1·96 | 2·02 (1·93-2·11) |
| Vaccine deficit | 2 | 616 | 42870253 | 5·24 | 5·00 (4·61-5·43) |
| Vaccine deficit | 3 | 1011 | 315608655 | 1·17 | 1·29 (1·21-1·38) |
| MCS class | 0 | 4798 | 2205786333 | 0·79 | Ref |
| MCS class | 1-4 | 4322 | 952682589 | 1·66 | 2·10 (2·01-2·19) |
| MCS class | 5-9 | 793 | 106445033 | 2·72 | 3·61 (3·34-3·89) |
| MCS class | 10-14 | 521 | 57159294 | 3·33 | 4·18 (3·82-4·58) |
| MCS class | 15-19 | 281 | 33725496 | 3·04 | 3·78 (3·35-4·26) |
| MCS class | >20 | 440 | 26039928 | 6·17 | 8·37 (7·59-9·24) |
| Sex | F | 6086 | 1687356955 | 1·32 | Ref |
| Sex | M | 5069 | 1694481718 | 1·09 | 0·92 (0·89-0·96) |
| Urbanisation class | Rural | 15 | 4666911 | 1·17 | 0·81 (0·49-1·35) |
| Urbanisation class | Suburban | 4304 | 1522934815 | 1·03 | 0·77 (0·75-0·80) |
| Urbanisation class | Urban | 6836 | 1854236947 | 1·35 | Ref |

Table S9: Basic adjustment HR - under-60 long COVID-19 analysis. Number of events, person-time (days), event rate per 1,000 individuals per year, adjusted HR with 95% confidence intervals.

##### Over 60 – Basic adjustment coefficients plots

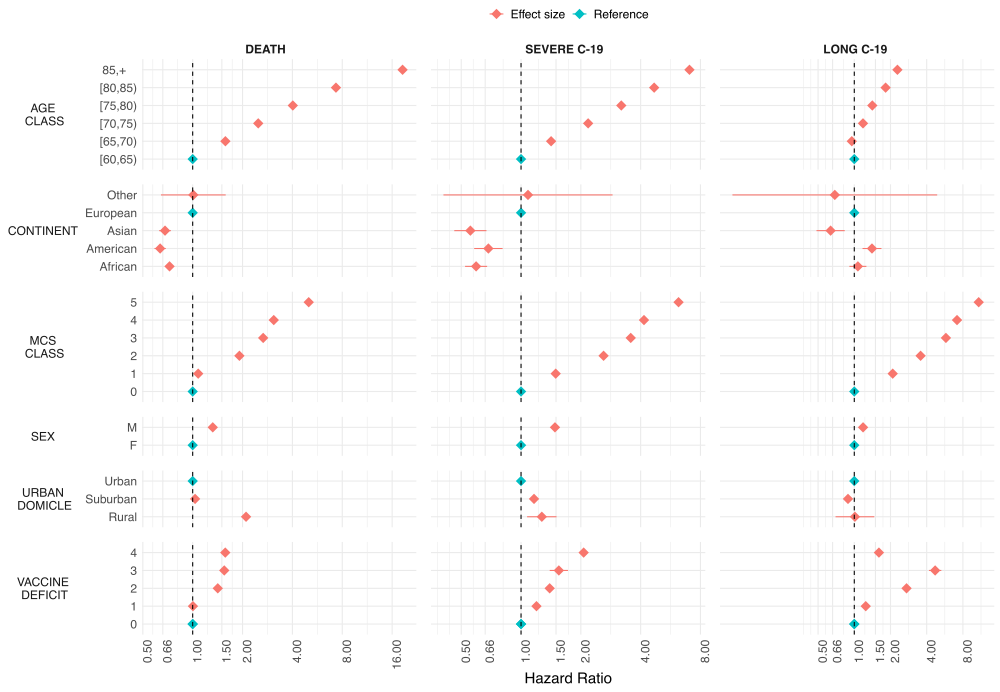

Figure S5: Forest plot basic adjustment analysis – over-60

##### Over 60 - Death outcome coefficients

| **Variable** | **Level** | **Events** | **Person time** | **Annual event rate**  **x1000 individuals** | **Adjusted HR**  **(95% CI)** |
| --- | --- | --- | --- | --- | --- |
| Age class | [60,65) | 10107 | 602832461 | 6·12 | Ref |
| Age class | [65,70) | 14471 | 516191233 | 10·23 | 1·58 (1·54-1·62) |
| Age class | [70,75) | 23758 | 497018700 | 17·45 | 2·49 (2·43-2·55) |
| Age class | [75,80) | 32582 | 390936974 | 30·42 | 4·02 (3·93-4·11) |
| Age class | [80,85) | 53798 | 332872446 | 58·99 | 7·33 (7·17-7·49) |
| Age class | 85+ | 113101 | 262585235 | 157·21 | 18·50 (18·11-18·89) |
| Continent of birth | African | 1366 | 29920515 | 16·66 | 0·72 (0·69-0·76) |
| Continent of birth | American | 624 | 23707470 | 9·61 | 0·64 (0·59-0·69) |
| Continent of birth | Asian | 583 | 21951257 | 9·69 | 0·68 (0·63-0·74) |
| Continent of birth | European | 245225 | 2526449669 | 35·43 | Ref |
| Continent of birth | Other | 19 | 408138 | 16·99 | 1·01 (0·64-1·58) |
| Vaccine deficit | 0 | 112691 | 925776939 | 44·43 | Ref |
| Vaccine deficit | 1 | 99302 | 1328636664 | 27·28 | 1·00 (0·99-1·01) |
| Vaccine deficit | 2 | 14931 | 166818444 | 32·67 | 1·42 (1·39-1·44) |
| Vaccine deficit | 3 | 2054 | 17859285 | 41·98 | 1·55 (1·48-1·62) |
| Vaccine deficit | 4 | 18839 | 163345717 | 42·10 | 1·57 (1·55-1·60) |
| MCS class | 0 | 40315 | 944583297 | 15·58 | Ref |
| MCS class | 1-4 | 53896 | 958730353 | 20·52 | 1·08 (1·07-1·09) |
| MCS class | 5-9 | 54335 | 361869625 | 54·81 | 1·91 (1·89-1·94) |
| MCS class | 10-14 | 27343 | 136243204 | 73·25 | 2·66 (2·62-2·71) |
| MCS class | 15-19 | 17842 | 75578524 | 86·17 | 3·09 (3·03-3·14) |
| MCS class | >20 | 54086 | 125432046 | 157·39 | 5·02 (4·95-5·08) |
| Sex | F | 133490 | 1443223943 | 33·76 | Ref |
| Sex | M | 114327 | 1159213106 | 36·00 | 1·32 (1·31-1·33) |
| Urbanisation class | Rural | 1485 | 4683138 | 115·74 | 2·10 (1·99-2·21) |
| Urbanisation class | Suburban | 108567 | 1159971160 | 34·16 | 1·03 (1·03-1·04) |
| Urbanisation class | Urban | 137765 | 1437782751 | 34·97 | Ref |

Table S10: Basic adjustment HR - over-60 mortality analysis. Number of events, person-time (days), event rate per 1,000 individuals per year, adjusted HR with 95% confidence intervals.

##### Over 60 - Severe COVID-19 outcome coefficients

| **Variable** | **Level** | **Events** | **Person time** | **Annual event rate**  **x1000 individuals** | **Adjusted HR**  **(95% CI)** |
| --- | --- | --- | --- | --- | --- |
| Age class | [60,65) | 3839 | 514474740 | 2·72 | Ref |
| Age class | [65,70) | 2927 | 601446400 | 1·78 | 1·42 (1·35-1·49) |
| Age class | [70,75) | 6224 | 494262103 | 4·60 | 2·18 (2·08-2·28) |
| Age class | [75,80) | 7792 | 387731057 | 7·34 | 3·20 (3·06-3·34) |
| Age class | [80,85) | 10304 | 328895690 | 11·44 | 4·68 (4·49-4·88) |
| Age class | 85+ | 13255 | 258502342 | 18·72 | 7·04 (6·76-7·34) |
| Continent of birth | African | 238 | 29818100 | 2·91 | 0·59 (0·52-0·67) |
| Continent of birth | American | 143 | 23646492 | 2·21 | 0·68 (0·58-0·81) |
| Continent of birth | Asian | 111 | 21900734 | 1·85 | 0·56 (0·46-0·67) |
| Continent of birth | European | 43845 | 2509540245 | 6·38 | Ref |
| Continent of birth | Other | 4 | 406761 | 3·59 | 1·08 (0·41-2·89) |
| Vaccine deficit | 0 | 15593 | 919303377 | 6·19 | Ref |
| Vaccine deficit | 1 | 21954 | 1320437166 | 6·07 | 1·20 (1·17-1·22) |
| Vaccine deficit | 2 | 2622 | 165830935 | 5·77 | 1·39 (1·34-1·45) |
| Vaccine deficit | 3 | 351 | 17728679 | 7·23 | 1·55 (1·39-1·72) |
| Vaccine deficit | 4 | 3821 | 162012175 | 8·61 | 2·07 (1·99-2·14) |
| MCS class | 0 | 6492 | 941881512 | 2·52 | Ref |
| MCS class | 1-4 | 11132 | 953992106 | 4·26 | 1·49 (1·45-1·54) |
| MCS class | 5-9 | 9655 | 358175281 | 9·84 | 2·60 (2·52-2·69) |
| MCS class | 10-14 | 4930 | 134424988 | 13·39 | 3·57 (3·43-3·70) |
| MCS class | 15-19 | 3249 | 74418430 | 15·94 | 4·16 (3·98-4·34) |
| MCS class | >20 | 8883 | 122420015 | 26·49 | 6·21 (6·01-6·42) |
| Sex | F | 21542 | 1434702161 | 5·48 | Ref |
| Sex | M | 22799 | 1150610171 | 7·23 | 1·48 (1·45-1·51) |
| Urbanisation class | Rural | 23119 | 1429006428 | 5·91 | 1·27 (1·07-1·51) |
| Urbanisation class | Suburban | 21089 | 1151665472 | 6·68 | 1·16 (1·14-1·18) |
| Urbanisation class | Urban | 133 | 4640432 | 10·46 | Ref |

Table S11: Basic adjustment HR - over-60 severe COVID-19 analysis. Number of events, person-time (days), event rate per 1,000 individuals per year, adjusted HR with 95% confidence intervals.

##### Over 60 - Long COVID-19 outcome coefficients

| **Variable** | **Level** | **Events** | **Person time** | **Annual event rate**  **x1000 individuals** | **Adjusted HR**  **(95% CI)** |
| --- | --- | --- | --- | --- | --- |
| Age class | [60,65) | 1843 | 570583215 | 1·18 | Ref |
| Age class | [65,70) | 1619 | 487404307 | 1·21 | 0·95 (0·89-1·02) |
| Age class | [70,75) | 2107 | 465154194 | 1·65 | 1·18 (1·11-1·26) |
| Age class | [75,80) | 2143 | 359752141 | 2·17 | 1·41 (1·33-1·51) |
| Age class | [80,85) | 2422 | 298945955 | 2·96 | 1·82 (1·71-1·94) |
| Age class | 85+ | 2491 | 227091999 | 4·00 | 2·28 (2·14-2·43) |
| Continent of birth | African | 148 | 27306247 | 1·98 | 1·07 (0·91-1·26) |
| Continent of birth | American | 120 | 22239963 | 1·97 | 1·40 (1·17-1·68) |
| Continent of birth | Asian | 53 | 20803973 | 0·93 | 0·64 (0·48-0·83) |
| Continent of birth | European | 12303 | 2338187053 | 1·92 | Ref |
| Continent of birth | Other | 1 | 394575 | 0·93 | 0·69 (0·10-4·89) |
| Vaccine deficit | 0 | 4338 | 850908951 | 1·86 | Ref |
| Vaccine deficit | 1 | 5751 | 1235891917 | 1·70 | 1·25 (1·20-1·30) |
| Vaccine deficit | 2 | 1438 | 153868731 | 3·41 | 2·72 (2·56-2·89) |
| Vaccine deficit | 3 | 295 | 16046497 | 6·71 | 4·70 (4·18-5·29) |
| Vaccine deficit | 4 | 803 | 152215715 | 1·93 | 1·60 (1·48-1·73) |
| MCS class | 0 | 1714 | 907342896 | 0·69 | Ref |
| MCS class | 1-4 | 3607 | 896144497 | 1·47 | 2·09 (1·97-2·21) |
| MCS class | 5-9 | 2462 | 323560931 | 2·78 | 3·56 (3·34-3·79) |
| MCS class | 10-14 | 1464 | 118855099 | 4·50 | 5·78 (5·38-6·20) |
| MCS class | 15-19 | 988 | 64143253 | 5·62 | 7·15 (6·60-7·74) |
| MCS class | >20 | 2390 | 98885135 | 8·82 | 10·83 (10·15-11·54) |
| Sex | F | 6536 | 1332274705 | 1·79 | Ref |
| Sex | M | 6089 | 1076657106 | 2·06 | 1·18 (1·14-1·23) |
| Urbanisation class | Rural | 28 | 4081073 | 2·50 | 1·01 (0·70-1·47) |
| Urbanisation class | Suburban | 5173 | 1066047681 | 1·77 | 0·89 (0·86-0·92) |
| Urbanisation class | Urban | 7424 | 1338803057 | 2·02 | Ref |

Table S12: Basic adjustment HR - over-60 long COVID-19 analysis. Number of events, person-time (days), event rate per 1,000 individuals per year, adjusted HR with 95% confidence intervals.

#### Full adjustment analysis

##### Under 60 – Full adjustment coefficients plots

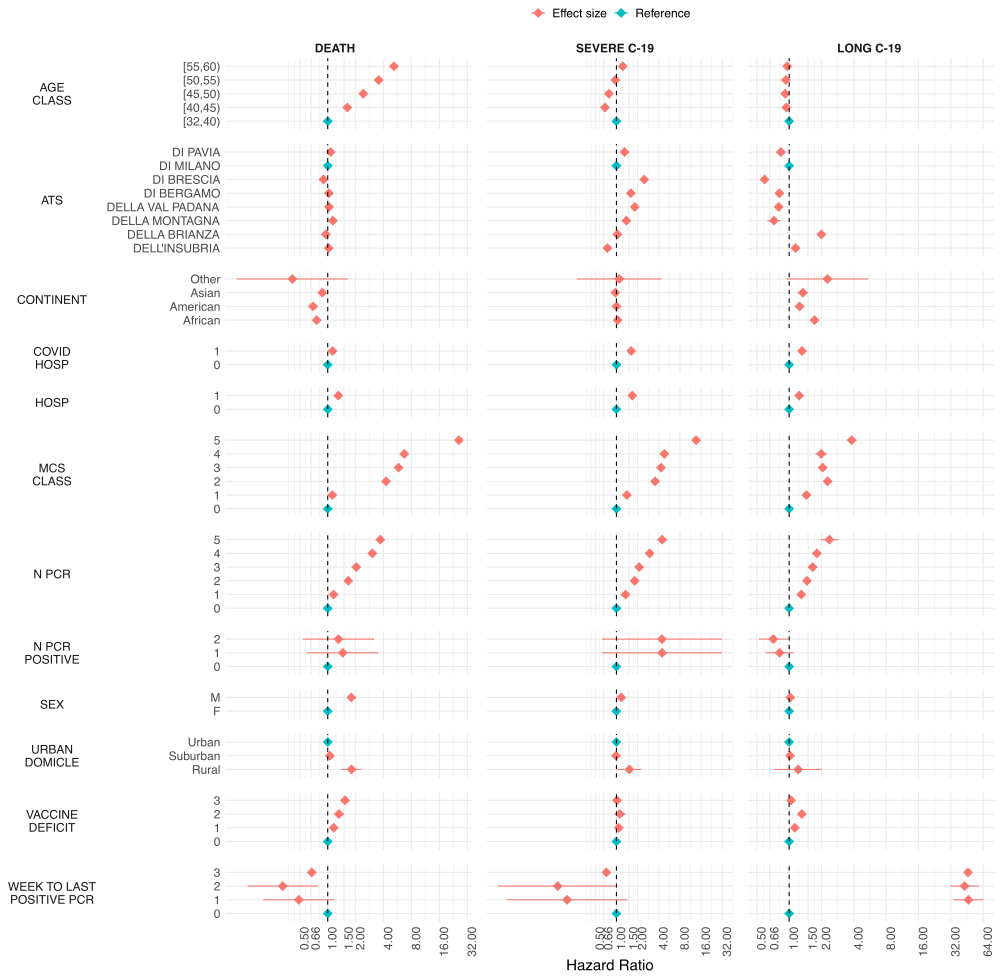

Figure S6: Forest plot full adjustment analysis – under-60..

##### Under 60 - Death outcome coefficients

| **Variable** | **Level** | **Events** | **Person time** | **Annual event rate**  **x1000 individuals** | **Adjusted HR**  **(95% CI)** |
| --- | --- | --- | --- | --- | --- |
| Age class | [32,40) | 1062 | 756792922 | 0·51 | Ref |
| Age class | [40,45) | 1376 | 583071906 | 0·86 | 1·63 (1·50-1·76) |
| Age class | [45,50) | 2733 | 720749661 | 1·38 | 2·42 (2·25-2·60) |
| Age class | [50,55) | 4658 | 753550030 | 2·26 | 3·54 (3·31-3·79) |
| Age class | [55,60) | 7596 | 734385807 | 3·78 | 5·18 (4·85-5·53) |
| ATS | ATS DELL'INSUBRIA | 2544 | 511212978 | 1·82 | 1·02 (0·98-1·07) |
| ATS | ATS DELLA BRIANZA | 1893 | 427564249 | 1·62 | 0·95 (0·90-1·00) |
| ATS | ATS DI MILANO | 5896 | 1248933524 | 1·72 | Ref |
| ATS | ATS DELLA MONTAGNA | 625 | 100902764 | 2·26 | 1·14 (1·04-1·24) |
| ATS | ATS DELLA VAL PADANA | 1435 | 266029184 | 1·97 | 1·03 (0·97-1·10) |
| ATS | ATS DI BERGAMO | 1973 | 393855965 | 1·83 | 1·03 (0·98-1·09) |
| ATS | ATS DI BRESCIA | 1902 | 413874525 | 1·68 | 0·90 (0·85-0·95) |
| ATS | ATS DI PAVIA | 1157 | 186177137 | 2·27 | 1·07 (1·01-1·15) |
| Covid Hospitalisations | 0 | 16642 | 3507385690 | 1·73 | Ref |
| Covid Hospitalisations | ≥ 1 | 783 | 41164636 | 6·94 | 1·13 (1·04-1·23) |
| Continent of birth | African | 565 | 153150282 | 1·35 | 0·76 (0·70-0·83) |
| Continent of birth | American | 280 | 101485617 | 1·01 | 0·69 (0·62-0·78) |
| Continent of birth | Asian | 457 | 131508449 | 1·27 | 0·88 (0·80-0·96) |
| Continent of birth | European | 16121 | 3161521744 | 1·86 | Ref |
| Continent of birth | Other | 2 | 884234 | 0·83 | 0·42 (0·10-1·66) |
| Vaccine deficit | 0 | 12947 | 2658942109 | 1·78 | Ref |
| Vaccine deficit | 1 | 2383 | 513412802 | 1·69 | 1·16 (1·11-1·21) |
| Vaccine deficit | 2 | 252 | 46310337 | 1·99 | 1·32 (1·17-1·50) |
| Vaccine deficit | 3 | 1843 | 329885078 | 2·04 | 1·54 (1·46-1·61) |
| Hospitalisations | 0 | 5916 | 2033771718 | 1·06 | 1·00 (1·00-1·00) |
| Hospitalisations | ≥ 1 | 11509 | 1514778608 | 2·77 | 1·30 (1·25-1·35) |
| Weeks to last PCR | None | 14670 | 2958058468 | 1·81 | Ref |
| Weeks to last PCR | 0-13 | 399 | 47444269 | 3·07 | 0·49 (0·20-1·18) |
| Weeks to last PCR | 14-26 | 927 | 227861748 | 1·48 | 0·33 (0·14-0·79) |
| Weeks to last PCR | ≥ 27 | 1429 | 315185841 | 1·65 | 0·67 (0·63-0·72) |
| MCS class | 0 | 6101 | 2276776799 | 0·98 | Ref |
| MCS class | 1-4 | 3317 | 1018910027 | 1·19 | 1·12 (1·07-1·17) |
| MCS class | 5-9 | 1800 | 118770703 | 5·53 | 4·26 (4·03-4·50) |
| MCS class | 10-14 | 1315 | 64126978 | 7·48 | 5·79 (5·43-6·18) |
| MCS class | 15-19 | 898 | 38150603 | 8·59 | 6·68 (6·20-7·21) |
| MCS class | >20 | 3994 | 31815216 | 45·82 | 25·97 (24·70-27·31) |
| Number of PCR | 0 | 12139 | 2864065614 | 1·55 | Ref |
| Number of PCR | 1 | 2096 | 412482399 | 1·85 | 1·16 (1·10-1·21) |
| Number of PCR | 2 | 1046 | 157372185 | 2·43 | 1·66 (1·56-1·78) |
| Number of PCR | 3 | 563 | 60861762 | 3·38 | 2·03 (1·85-2·22) |
| Number of PCR | 4-9 | 1244 | 48162079 | 9·43 | 3·03 (2·84-3·23) |
| Number of PCR | ≥10 | 337 | 5606287 | 21·94 | 3·69 (3·29-4·14) |
| Number of positive PCR | 0 | 16104 | 3275547082 | 1·79 | Ref |
| Number of positive PCR | 1 | 752 | 199039846 | 1·38 | 1·46 (0·60-3·51) |
| Number of positive PCR | ≥2 | 569 | 73963398 | 2·81 | 1·31 (0·54-3·16) |
| Sex | F | 6548 | 1782370848 | 1·34 | Ref |
| Sex | M | 10877 | 1766179478 | 2·25 | 1·80 (1·74-1·85) |
| Urban class | Rural | 64 | 4926845 | 4·74 | 1·81 (1·41-2·31) |
| Urban class | Suburban | 9114 | 1934710208 | 1·72 | 1·05 (1·02-1·09) |
| Urban class | Urban | 8247 | 1608913273 | 1·87 | Ref |

Table S13: Full adjustment HR - under-60 mortality analysis. Number of events, person-time (days), event rate per 1,000 individuals per year, adjusted HR with 95% confidence intervals.

##### Under 60 - Severe COVID-19 outcome coefficients

| **Variable** | **Level** | **Events** | **Person time** | **Annual event rate**  **x1000 individuals** | **Adjusted HR**  **(95% CI)** |
| --- | --- | --- | --- | --- | --- |
| Age class | [32,40) | 1574 | 755783862 | 0·76 | Ref |
| Age class | [40,45) | 850 | 582559074 | 0·53 | 0·69 (0·63-0·75) |
| Age class | [45,50) | 1226 | 720062773 | 0·62 | 0·78 (0·73-0·84) |
| Age class | [50,55) | 1698 | 752625535 | 0·82 | 0·96 (0·90-1·03) |
| Age class | [55,60) | 2344 | 733196285 | 1·17 | 1·23 (1·15-1·31) |
| ATS | ATS DELL'INSUBRIA | 631 | 510871558 | 0·45 | 0·75 (0·68-0·82) |
| ATS | ATS DELLA BRIANZA | 711 | 427191258 | 0·61 | 1·03 (0·95-1·13) |
| ATS | ATS DI MILANO | 2057 | 1247813829 | 0·60 | Ref |
| ATS | ATS DELLA MONTAGNA | 240 | 100771527 | 0·87 | 1·39 (1·21-1·59) |
| ATS | ATS DELLA VAL PADANA | 830 | 265572998 | 1·14 | 1·82 (1·67-1·99) |
| ATS | ATS DI BERGAMO | 1016 | 393247952 | 0·94 | 1·60 (1·48-1·74) |
| ATS | ATS DI BRESCIA | 1767 | 412792251 | 1·56 | 2·49 (2·32-2·66) |
| ATS | ATS DI PAVIA | 440 | 185966156 | 0·86 | 1·31 (1·18-1·45) |
| Covid Hospitalisations | 0 | 7275 | 3503267424 | 0·76 | Ref |
| Covid Hospitalisations | ≥ 1 | 417 | 40960105 | 3·72 | 1·62 (1·45-1·83) |
| Continent of birth | African | 367 | 152939251 | 0·88 | 1·04 (0·94-1·16) |
| Continent of birth | American | 190 | 101373052 | 0·68 | 1·00 (0·87-1·16) |
| Continent of birth | Asian | 262 | 131357351 | 0·73 | 0·97 (0·85-1·09) |
| Continent of birth | European | 6871 | 3157674927 | 0·79 | Ref |
| Continent of birth | Other | 2 | 882948 | 0·83 | 1·10 (0·27-4·39) |
| Vaccine deficit | 0 | 5783 | 2655718691 | 0·79 | Ref |
| Vaccine deficit | 1 | 1167 | 512744803 | 0·83 | 1·08 (1·01-1·15) |
| Vaccine deficit | 2 | 116 | 46244914 | 0·92 | 1·12 (0·93-1·35) |
| Vaccine deficit | 3 | 626 | 329519121 | 0·69 | 1·02 (0·94-1·11) |
| Hospitalisations | 0 | 2413 | 2032401866 | 0·43 | Ref |
| Hospitalisations | ≥ 1 | 5279 | 1511825663 | 1·27 | 1·68 (1·59-1·78) |
| Weeks to last PCR | None | 6188 | 2954547561 | 0·76 | Ref |
| Weeks to last PCR | 0-13 | 232 | 47324409 | 1·79 | 0·20 (0·03-1·42) |
| Weeks to last PCR | 14-26 | 596 | 227533832 | 0·96 | 0·15 (0·02-1·04) |
| Weeks to last PCR | ≥ 27 | 676 | 314821727 | 0·78 | 0·72 (0·66-0·79) |
| MCS class | 0 | 2888 | 2275067220 | 0·46 | 1·00 (1·00-1·00) |
| MCS class | 1-4 | 2100 | 1017689150 | 0·75 | 1·41 (1·33-1·49) |
| MCS class | 5-9 | 728 | 118378440 | 2·24 | 3·54 (3·25-3·86) |
| MCS class | 10-14 | 536 | 63838846 | 3·06 | 4·30 (3·90-4·74) |
| MCS class | 15-19 | 365 | 37952794 | 3·51 | 4·80 (4·28-5·38) |
| MCS class | >20 | 1075 | 31301079 | 12·54 | 13·56 (12·49-14·72) |
| Number of PCR | 0 | 5106 | 2861179855 | 0·65 | Ref |
| Number of PCR | 1 | 1105 | 411839300 | 0·98 | 1·35 (1·26-1·44) |
| Number of PCR | 2 | 539 | 157073658 | 1·25 | 1·82 (1·65-2·00) |
| Number of PCR | 3 | 276 | 60707058 | 1·66 | 2·10 (1·84-2·40) |
| Number of PCR | 4-9 | 502 | 47902079 | 3·83 | 2·97 (2·67-3·29) |
| Number of PCR | ≥10 | 164 | 5525579 | 10·83 | 4·47 (3·77-5·29) |
| Number of positive PCR | 0 | 6865 | 3271671351 | 0·77 | Ref |
| Number of positive PCR | 1 | 482 | 198775142 | 0·89 | 4·46 (0·63-31·74) |
| Number of positive PCR | ≥2 | 345 | 73781036 | 1·71 | 4·43 (0·62-31·61) |
| Sex | F | 3834 | 1780129296 | 0·79 | Ref |
| Sex | M | 3858 | 1764098233 | 0·80 | 1·17 (1·12-1·23) |
| Urban class | Rural | 25 | 4911190 | 1·86 | 1·53 (1·03-2·27) |
| Urban class | Suburban | 4059 | 1606630336 | 0·92 | 0·99 (0·94-1·04) |
| Urban class | Urban | 3608 | 1932686003 | 0·68 | Ref |

Table S14: Full adjustment HR - under-60 severe COVID-19 analysis. Number of events, person-time (days), event rate per 1,000 individuals per year, adjusted HR with 95% confidence intervals.

##### Under 60 - Long COVID-19 outcome coefficients

| **Variable** | **Level** | **Events** | **Person time** | **Annual event rate**  **x1000 individuals** | **Adjusted HR**  **(95% CI)** |
| --- | --- | --- | --- | --- | --- |
| Age class | [32,40) | 2608 | 712037437 | 1·34 | Ref |
| Age class | [40,45) | 1835 | 549227708 | 1·22 | 0·94 (0·89-1·00) |
| Age class | [45,50) | 2146 | 680968001 | 1·15 | 0·92 (0·87-0·97) |
| Age class | [50,55) | 2246 | 713749430 | 1·15 | 0·93 (0·88-0·99) |
| Age class | [55,60) | 2223 | 696584783 | 1·16 | 0·96 (0·90-1·02) |
| ATS | ATS DELL'INSUBRIA | 1934 | 469744595 | 1·50 | 1·15 (1·09-1·21) |
| ATS | ATS DELLA BRIANZA | 2476 | 387198164 | 2·33 | 1·99 (1·90-2·10) |
| ATS | ATS DI MILANO | 3893 | 1197147209 | 1·19 | Ref |
| ATS | ATS DELLA MONTAGNA | 212 | 96822919 | 0·80 | 0·72 (0·63-0·83) |
| ATS | ATS DELLA VAL PADANA | 639 | 246341281 | 0·95 | 0·80 (0·73-0·87) |
| ATS | ATS DI BERGAMO | 675 | 374789517 | 0·66 | 0·81 (0·75-0·88) |
| ATS | ATS DI BRESCIA | 795 | 400729126 | 0·72 | 0·59 (0·55-0·64) |
| ATS | ATS DI PAVIA | 434 | 179794548 | 0·88 | 0·84 (0·76-0·93) |
| Covid Hospitalisations | 0 | 9961 | 3317658686 | 1·10 | Ref |
| Covid Hospitalisations | ≥ 1 | 1097 | 34908673 | 11·47 | 1·32 (1·23-1·41) |
| Continent of birth | African | 536 | 139621470 | 1·40 | 1·72 (1·58-1·88) |
| Continent of birth | American | 482 | 95317697 | 1·85 | 1·25 (1·14-1·37) |
| Continent of birth | Asian | 345 | 124661425 | 1·01 | 1·34 (1·20-1·50) |
| Continent of birth | European | 9690 | 2992128957 | 1·18 | Ref |
| Continent of birth | Other | 5 | 837810 | 2·18 | 2·27 (0·95-5·46) |
| Vaccine deficit | 0 | 6867 | 2516345381 | 1·00 | Ref |
| Vaccine deficit | 1 | 2583 | 482288227 | 1·95 | 1·13 (1·07-1·19) |
| Vaccine deficit | 2 | 611 | 42344778 | 5·27 | 1·31 (1·21-1·43) |
| Vaccine deficit | 3 | 997 | 311588973 | 1·17 | 1·04 (0·97-1·12) |
| Hospitalisations | 0 | 4547 | 1954111858 | 0·85 | Ref |
| Hospitalisations | ≥ 1 | 6511 | 1398455501 | 1·70 | 1·24 (1·19-1·29) |
| Weeks to last PCR | None | 957 | 2807325212 | 0·12 | Ref |
| Weeks to last PCR | 0-13 | 869 | 43438997 | 7·30 | 46·71 (34·00-64·18) |
| Weeks to last PCR | 14-26 | 3757 | 210881976 | 6·50 | 42·97 (31·53-58·56) |
| Weeks to last PCR | ≥ 27 | 5475 | 290921174 | 6·87 | 46·25 (43·12-49·60) |
| MCS class | 0 | 4743 | 2182742093 | 0·79 | Ref |
| MCS class | 1-4 | 4297 | 947688237 | 1·65 | 1·45 (1·39-1·51) |
| MCS class | 5-9 | 789 | 105909909 | 2·72 | 2·28 (2·11-2·47) |
| MCS class | 10-14 | 516 | 56813015 | 3·32 | 2·05 (1·86-2·26) |
| MCS class | 15-19 | 281 | 33514169 | 3·06 | 1·99 (1·76-2·25) |
| MCS class | >20 | 432 | 25899936 | 6·09 | 3·82 (3·44-4·25) |
| Number of PCR | 0 | 4966 | 2719774747 | 0·67 | Ref |
| Number of PCR | 1 | 2494 | 383929688 | 2·37 | 1·30 (1·22-1·38) |
| Number of PCR | 2 | 1911 | 145119730 | 4·81 | 1·46 (1·36-1·58) |
| Number of PCR | 3 | 870 | 55577386 | 5·71 | 1·66 (1·51-1·82) |
| Number of PCR | 4-9 | 701 | 43209596 | 5·92 | 1·81 (1·64-2·00) |
| Number of PCR | ≥10 | 116 | 4956212 | 8·54 | 2·37 (1·95-2·88) |
| Number of positive PCR | 0 | 6475 | 3100402256 | 0·76 | Ref |
| Number of positive PCR | 1 | 3298 | 184275246 | 6·53 | 0·81 (0·60-1·10) |
| Number of positive PCR | ≥2 | 1285 | 67889857 | 6·91 | 0·71 (0·52-0·97) |
| Sex | F | 6053 | 1677327798 | 1·32 | Ref |
| Sex | M | 5005 | 1675239561 | 1·09 | 1·02 (0·98-1·06) |
| Urban class | Rural | 15 | 4617906 | 1·19 | 1·21 (0·73-2·02) |
| Urban class | Suburban | 4274 | 1510809747 | 1·03 | 1·02 (0·98-1·06) |
| Urban class | Urban | 6769 | 1837139706 | 1·34 | Ref |

Table S15: Full adjustment HR - under-60 long COVID-19 analysis. Number of events, person-time (days), event rate per 1,000 individuals per year, adjusted HR with 95% confidence intervals.

##### Over 60 – Full adjustment coefficients plots

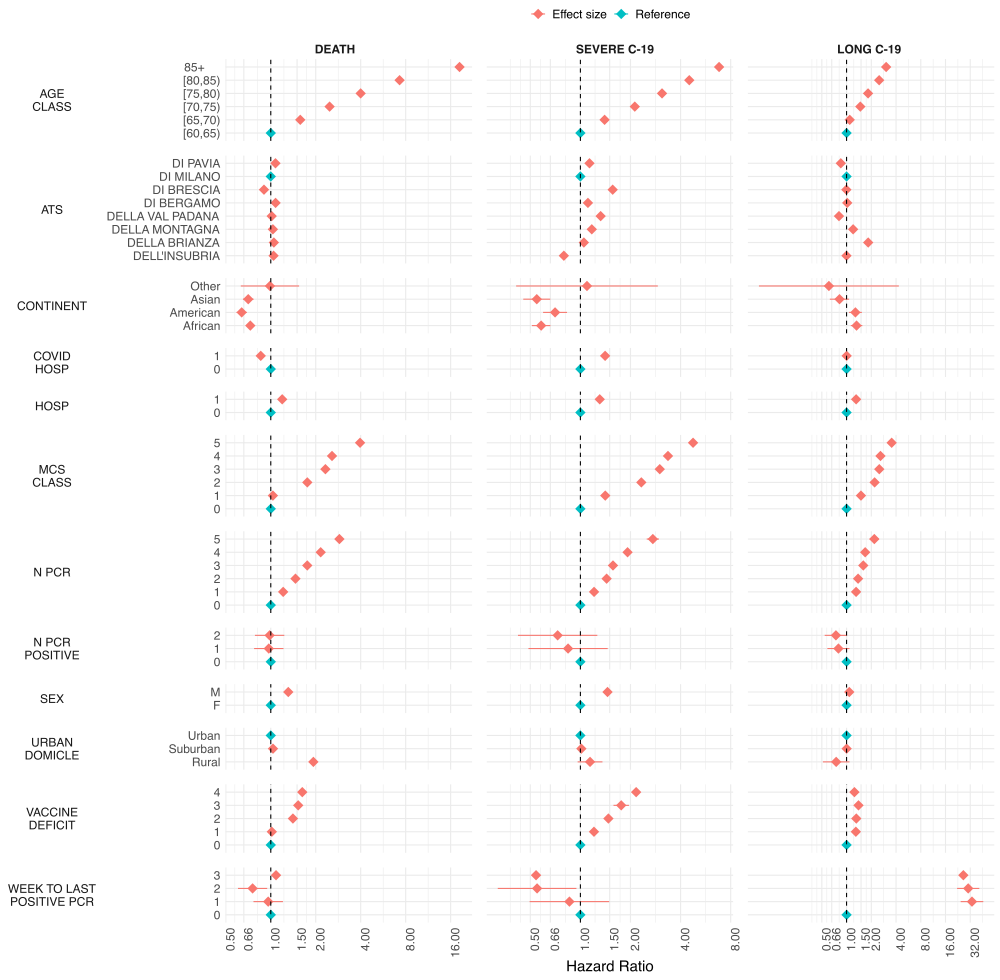

Figure S7: Forest plot full adjustment analysis – over-60

##### Over 60 - Death outcome coefficients

| **Variable** | **Level** | **Events** | **Person time** | **Annual event rate**  **x1000 individuals** | **Adjusted HR**  **(95% CI)** |
| --- | --- | --- | --- | --- | --- |
| Age class | [60,65) | 10048 | 600444577 | 6·11 | Ref |
| Age class | [65,70) | 14413 | 514653643 | 10·22 | 1·58 (1·54-1·62) |
| Age class | [70,75) | 23689 | 496004142 | 17·43 | 2·48 (2·42-2·53) |
| Age class | [75,80) | 32522 | 390274389 | 30·42 | 4·00 (3·91-4·09) |
| Age class | [80,85) | 53710 | 332388975 | 58·98 | 7·28 (7·13-7·44) |
| Age class | 85+ | 112933 | 262197173 | 157·21 | 18·36 (17·98-18·75) |
| ATS | ATS DELL'INSUBRIA | 36964 | 387511623 | 34·82 | 1·04 (1·03-1·06) |
| ATS | ATS DELLA BRIANZA | 29096 | 318336007 | 33·36 | 1·05 (1·03-1·06) |
| ATS | ATS DI MILANO | 84902 | 891542187 | 34·76 | Ref |
| ATS | ATS DELLA MONTAGNA | 7642 | 79974338 | 34·88 | 1·03 (1·01-1·06) |
| ATS | ATS DELLA VAL PADANA | 21139 | 206743482 | 37·32 | 1·01 (1·00-1·03) |
| ATS | ATS DI BERGAMO | 25201 | 274835890 | 33·47 | 1·08 (1·06-1·09) |
| ATS | ATS DI BRESCIA | 25963 | 289970537 | 32·68 | 0·90 (0·89-0·91) |
| ATS | ATS DI PAVIA | 16408 | 147048835 | 40·73 | 1·07 (1·06-1·09) |
| Covid Hospitalisations | 0 | 232845 | 2528870266 | 33·61 | Ref |
| Covid Hospitalisations | ≥ 1 | 14470 | 67092633 | 78·72 | 0·86 (0·84-0·87) |
| Continent of birth | African | 1362 | 29722481 | 16·73 | 0·73 (0·69-0·77) |
| Continent of birth | American | 620 | 23544138 | 9·61 | 0·64 (0·59-0·69) |
| Continent of birth | Asian | 582 | 21830870 | 9·73 | 0·71 (0·65-0·77) |
| Continent of birth | European | 244732 | 2520458216 | 35·44 | Ref |
| Continent of birth | Other | 19 | 407194 | 17·03 | 0·99 (0·63-1·55) |
| Vaccine deficit | 0 | 112508 | 924352407 | 44·43 | Ref |
| Vaccine deficit | 1 | 99144 | 1325523071 | 27·30 | 1·02 (1·01-1·03) |
| Vaccine deficit | 2 | 14869 | 165984539 | 32·70 | 1·41 (1·38-1·43) |
| Vaccine deficit | 3 | 2047 | 17766289 | 42·05 | 1·53 (1·46-1·60) |
| Vaccine deficit | 4 | 18747 | 162336593 | 42·15 | 1·63 (1·60-1·65) |
| Hospitalisations | 0 | 55555 | 1125592793 | 18·02 | Ref |
| Hospitalisations | ≥ 1 | 191760 | 1470370106 | 47·60 | 1·19 (1·18-1·21) |
| Weeks to last PCR | None | 211042 | 2319211711 | 33·21 | Ref |
| Weeks to last PCR | 0-13 | 5922 | 27778094 | 77·81 | 0·96 (0·77-1·21) |
| Weeks to last PCR | 14-26 | 9744 | 77972579 | 45·61 | 0·76 (0·60-0·95) |
| Weeks to last PCR | ≥ 27 | 20607 | 171000515 | 43·99 | 1·08 (1·07-1·10) |
| MCS class | 0 | 40148 | 940876361 | 15·57 | Ref |
| MCS class | 1-4 | 53811 | 957081955 | 20·52 | 1·03 (1·02-1·05) |
| MCS class | 5-9 | 54257 | 361332358 | 54·81 | 1·76 (1·73-1·78) |
| MCS class | 10-14 | 27301 | 135998358 | 73·27 | 2·32 (2·28-2·36) |
| MCS class | 15-19 | 17814 | 75460724 | 86·17 | 2·57 (2·52-2·62) |
| MCS class | >20 | 53984 | 125213143 | 157·36 | 3·96 (3·90-4·02) |
| Number of PCR | 0 | 183179 | 2217123327 | 30·16 | Ref |
| Number of PCR | 1 | 25001 | 220940493 | 41·30 | 1·21 (1·19-1·23) |
| Number of PCR | 2 | 12135 | 78587173 | 56·36 | 1·46 (1·44-1·49) |
| Number of PCR | 3 | 7888 | 34317182 | 83·90 | 1·76 (1·71-1·80) |
| Number of PCR | 4-9 | 15430 | 38881257 | 144·85 | 2·16 (2·12-2·20) |
| Number of PCR | ≥10 | 3682 | 6113467 | 219·83 | 2·88 (2·78-2·98) |
| Number of positive PCR | 0 | 231725 | 2491265368 | 33·95 | Ref |
| Number of positive PCR | 1 | 7122 | 67322207 | 38·61 | 0·97 (0·77-1·22) |
| Number of positive PCR | ≥2 | 8468 | 37375324 | 82·70 | 0·98 (0·78-1·23) |
| Sex | F | 133271 | 1440381255 | 33·77 | Ref |
| Sex | M | 114044 | 1155581644 | 36·02 | 1·31 (1·30-1·32) |
| Urban class | Rural | 1480 | 4664875 | 115·80 | 1·93 (1·83-2·03) |
| Urban class | Suburban | 108327 | 1157025696 | 34·17 | 1·03 (1·03-1·04) |
| Urban class | Urban | 137508 | 1434272328 | 34·99 | Ref |

Table S16: Full adjustment HR - over-60 mortality analysis. Number of events, person-time (days), event rate per 1,000 individuals per year, adjusted HR with 95% confidence intervals.

##### Over 60 - Severe COVID-19 outcome

| **Variable** | **Level** | **Events** | **Person time** | **Annual event rate**  **x1000 individuals** | **Adjusted HR**  **(95% CI)** |
| --- | --- | --- | --- | --- | --- |
| Age class | [60,65) | 2918 | 599062975 | 1·78 | Ref |
| Age class | [65,70) | 3823 | 512946934 | 2·72 | 1·40 (1·33-1·47) |
| Age class | [70,75) | 6209 | 493253807 | 4·59 | 2·12 (2·03-2·22) |
| Age class | [75,80) | 7777 | 387074121 | 7·33 | 3·09 (2·96-3·23) |
| Age class | [80,85) | 10284 | 328422195 | 11·43 | 4·51 (4·33-4·71) |
| Age class | 85+ | 13238 | 258120017 | 18·72 | 6·82 (6·54-7·11) |
| ATS | ATS DELL'INSUBRIA | 4670 | 385746326 | 4·42 | 0·80 (0·77-0·82) |
| ATS | ATS DELLA BRIANZA | 4901 | 316472180 | 5·65 | 1·05 (1·02-1·08) |
| ATS | ATS DI MILANO | 14017 | 886483596 | 5·77 | Ref |
| ATS | ATS DELLA MONTAGNA | 1432 | 79432238 | 6·58 | 1·17 (1·11-1·24) |
| ATS | ATS DELLA VAL PADANA | 4500 | 205012519 | 8·01 | 1·32 (1·28-1·37) |
| ATS | ATS DI BERGAMO | 4530 | 272935748 | 6·06 | 1·11 (1·07-1·15) |
| ATS | ATS DI BRESCIA | 7361 | 286741068 | 9·37 | 1·56 (1·52-1·61) |
| ATS | ATS DI PAVIA | 2838 | 146056374 | 7·09 | 1·13 (1·09-1·18) |
| Covid Hospitalisations | 0 | 41661 | 2512657319 | 6·05 | Ref |
| Covid Hospitalisations | ≥ 1 | 2588 | 66222730 | 14·26 | 1·41 (1·34-1·48) |
| Continent of birth | African | 237 | 29620517 | 2·92 | 0·58 (0·51-0·66) |
| Continent of birth | American | 141 | 23484628 | 2·19 | 0·70 (0·60-0·83) |
| Continent of birth | Asian | 111 | 21780347 | 1·86 | 0·55 (0·45-0·66) |
| Continent of birth | European | 43756 | 2503588740 | 6·38 | Ref |
| Continent of birth | Other | 4 | 405817 | 3·60 | 1·10 (0·41-2·92) |
| Vaccine deficit | 0 | 15575 | 917889540 | 6·19 | Ref |
| Vaccine deficit | 1 | 21910 | 1317341701 | 6·07 | 1·21 (1·18-1·23) |
| Vaccine deficit | 2 | 2614 | 165000772 | 5·78 | 1·47 (1·41-1·54) |
| Vaccine deficit | 3 | 349 | 17636912 | 7·22 | 1·76 (1·58-1·96) |
| Vaccine deficit | 4 | 3801 | 161011124 | 8·62 | 2·16 (2·09-2·24) |
| Hospitalisations | 0 | 9487 | 1121749935 | 3·09 | Ref |
| Hospitalisations | ≥ 1 | 34762 | 1457130114 | 8·71 | 1·31 (1·24-1·37) |
| Weeks to last PCR | None | 39616 | 2303764119 | 6·28 | Ref |
| Weeks to last PCR | 0-13 | 916 | 27474498 | 12·17 | 0·86 (0·50-1·49) |
| Weeks to last PCR | 14-26 | 1224 | 77513802 | 5·76 | 0·55 (0·32-0·95) |
| Weeks to last PCR | ≥ 27 | 2493 | 170127630 | 5·35 | 0·54 (0·52-0·57) |
| MCS class | 0 | 6461 | 938190773 | 2·51 | Ref |
| MCS class | 1-4 | 11110 | 952352314 | 4·26 | 1·41 (1·37-1·46) |
| MCS class | 5-9 | 9644 | 357642715 | 9·84 | 2·32 (2·25-2·40) |
| MCS class | 10-14 | 4925 | 134181841 | 13·40 | 3·00 (2·88-3·12) |
| MCS class | 15-19 | 3245 | 74302310 | 15·94 | 3·36 (3·21-3·52) |
| MCS class | >20 | 8864 | 122210096 | 26·47 | 4·75 (4·58-4·93) |
| Number of PCR | 0 | 33375 | 2204127185 | 5·53 | Ref |
| Number of PCR | 1 | 4658 | 219094692 | 7·76 | 1·21 (1·17-1·25) |
| Number of PCR | 2 | 2073 | 77807312 | 9·72 | 1·44 (1·37-1·51) |
| Number of PCR | 3 | 1176 | 33907681 | 12·66 | 1·57 (1·48-1·67) |
| Number of PCR | 4-9 | 2340 | 38049485 | 22·45 | 1·92 (1·83-2·01) |
| Number of PCR | ≥10 | 627 | 5893694 | 38·83 | 2·73 (2·50-2·97) |
| Number of positive PCR | 0 | 1014 | 66947626 | 5·53 | Ref |
| Number of positive PCR | 1 | 42122 | 2474938823 | 6·21 | 0·84 (0·49-1·46) |
| Number of positive PCR | ≥2 | 1113 | 36993600 | 10·98 | 0·73 (0·42-1·26) |
| Sex | F | 21503 | 1431876602 | 5·48 | Ref |
| Sex | M | 22746 | 1147003447 | 7·24 | 1·46 (1·43-1·48) |
| Urban class | Rural | 132 | 4622177 | 10·42 | 1·14 (0·96-1·36) |
| Urban class | Suburban | 21050 | 1148734836 | 6·69 | 1·01 (0·99-1·04) |
| Urban class | Urban | 23067 | 1425523036 | 5·91 | Ref |

Table S17: Full adjustment HR - over-60 severe COVID-19 analysis. Number of events, person-time (days), event rate per 1,000 individuals per year, adjusted HR with 95% confidence intervals.

##### Over 60 - Long COVID-19 outcome

| **Variable** | **Level** | **Events** | **Person time** | **Annual event rate**  **x1000 individuals** | **Adjusted HR**  **(95% CI)** |
| --- | --- | --- | --- | --- | --- |
| Age class | [60,65) | 1833 | 568329491 | 1·18 | Ref |
| Age class | [65,70) | 1616 | 485945513 | 1·21 | 1·09 (1·02-1·17) |
| Age class | [70,75) | 2102 | 464208782 | 1·65 | 1·47 (1·38-1·57) |
| Age class | [75,80) | 2142 | 359138345 | 2·18 | 1·83 (1·71-1·95) |
| Age class | [80,85) | 2418 | 298508231 | 2·96 | 2·49 (2·34-2·66) |
| Age class | 85+ | 2490 | 226745856 | 4·01 | 3·03 (2·84-3·23) |
| ATS | ATS DELL'INSUBRIA | 1831 | 349017596 | 1·91 | 1·00 (0·94-1·05) |
| ATS | ATS DELLA BRIANZA | 2379 | 283145182 | 3·07 | 1·83 (1·74-1·93) |
| ATS | ATS DI MILANO | 4224 | 837883059 | 1·84 | Ref |
| ATS | ATS DELLA MONTAGNA | 425 | 73960827 | 2·10 | 1·20 (1·08-1·33) |
| ATS | ATS DELLA VAL PADANA | 763 | 187885133 | 1·48 | 0·81 (0·75-0·88) |
| ATS | ATS DI BERGAMO | 925 | 256845686 | 1·31 | 1·02 (0·95-1·10) |
| ATS | ATS DI BRESCIA | 1483 | 274323837 | 1·97 | 0·99 (0·93-1·06) |
| ATS | ATS DI PAVIA | 571 | 139814898 | 1·49 | 0·85 (0·78-0·93) |
| Covid Hospitalisations | 0 | 9439 | 2347794960 | 1·47 | Ref |
| Covid Hospitalisations | ≥ 1 | 3162 | 55081258 | 20·95 | 1·00 (0·96-1·05) |
| Continent of birth | African | 148 | 27118726 | 1·99 | 1·32 (1·12-1·56) |
| Continent of birth | American | 119 | 22087333 | 1·97 | 1·28 (1·07-1·54) |
| Continent of birth | Asian | 53 | 20687362 | 0·94 | 0·82 (0·63-1·08) |
| Continent of birth | European | 12280 | 2332589166 | 1·92 | Ref |
| Continent of birth | Other | 1 | 393631 | 0·93 | 0·61 (0·09-4·33) |
| Vaccine deficit | 0 | 4333 | 849575665 | 1·86 | Ref |
| Vaccine deficit | 1 | 5742 | 1232984636 | 1·70 | 1·30 (1·24-1·35) |
| Vaccine deficit | 2 | 1432 | 153095980 | 3·41 | 1·31 (1·23-1·40) |
| Vaccine deficit | 3 | 293 | 15963082 | 6·70 | 1·40 (1·24-1·57) |
| Vaccine deficit | 4 | 801 | 151256855 | 1·93 | 1·25 (1·15-1·35) |
| Hospitalisations | 0 | 2595 | 1080399962 | 0·88 | Ref |
| Hospitalisations | ≥ 1 | 10006 | 1322476256 | 2·76 | 1·31 (1·24-1·37) |
| Weeks to last PCR | None | 2585 | 2159014866 | 0·44 | Ref |
| Weeks to last PCR | 0-13 | 1383 | 23732297 | 21·27 | 33·58 (24·44-46·14) |
| Weeks to last PCR | 14-26 | 2938 | 68892324 | 15·57 | 30·23 (22·14-41·28) |
| Weeks to last PCR | ≥ 27 | 5695 | 151236731 | 13·74 | 26·38 (25·09-27·74) |
| MCS class | 0 | 1708 | 903779216 | 0·69 | Ref |
| MCS class | 1-4 | 3603 | 894619234 | 1·47 | 1·50 (1·41-1·59) |
| MCS class | 5-9 | 2457 | 323073118 | 2·78 | 2·19 (2·06-2·34) |
| MCS class | 10-14 | 1462 | 118638254 | 4·50 | 2·50 (2·32-2·70) |
| MCS class | 15-19 | 988 | 64049317 | 5·63 | 2·59 (2·37-2·82) |
| MCS class | >20 | 2383 | 98717079 | 8·81 | 3·54 (3·29-3·80) |
| Number of PCR | 0 | 6501 | 2067877523 | 1·15 | Ref |
| Number of PCR | 1 | 2081 | 199405313 | 3·81 | 1·31 (1·23-1·39) |
| Number of PCR | 2 | 1489 | 69552439 | 7·81 | 1·38 (1·28-1·49) |
| Number of PCR | 3 | 927 | 29607750 | 11·43 | 1·60 (1·46-1·74) |
| Number of PCR | 4-9 | 1263 | 31786100 | 14·50 | 1·69 (1·55-1·83) |
| Number of PCR | ≥10 | 340 | 4647093 | 26·70 | 2·18 (1·92-2·47) |
| Number of positive PCR | 0 | 8321 | 2311200950 | 1·31 | 1·00 (1·00-1·00) |
| Number of positive PCR | 1 | 2474 | 59852938 | 15·09 | 0·80 (0·58-1·09) |
| Number of positive PCR | ≥2 | 1806 | 31822330 | 20·71 | 0·74 (0·54-1·02) |
| Sex | F | 6529 | 1329624353 | 1·79 | Ref |
| Sex | M | 6072 | 1073251865 | 2·07 | 1·08 (1·04-1·12) |
| Urban class | Rural | 28 | 4065242 | 2·51 | 0·75 (0·51-1·09) |
| Urban class | Suburban | 5165 | 1063315983 | 1·77 | 1·00 (0·96-1·04) |
| Urban class | Urban | 1833 | 568329491 | 1·18 | Ref |

Table S18: Full adjustment HR - over-60 long COVID-19 analysis. Number of events, person-time (days), event rate per 1,000 individuals per year, adjusted HR with 95% confidence intervals.

### Long-COVID definition

Long COVID was operationalised as a symptom-based outcome using diagnostic codes consistent with the WHO clinical case definition of post-COVID condition.

We considered symptom codes covering (non-exhaustive): fatigue/malaise, memory loss/cognitive symptoms, altered mental status, generalised pain, gait abnormality/coordination problems, dyspnoea, and related symptom codes

Data sources (where symptoms were captured). Eligible symptom codes were ascertained from linked administrative health records, including: hospital discharge records, emergency department contacts, home-care registries, and care-home registries. To minimise capture of pre-existing symptoms, only symptom diagnoses recorded ≥1 month after infection were considered.

| **long covid-19 symptom** | **period** | **events** | **events per year** |
| --- | --- | --- | --- |
| Malaise and fatigue (ICD9=7807) | 2012-2019 | 993 | 124 |
|  | 2020-2024 | 4 243 | 849 |
| Chronic fatigue syndrome (ICD9=78071) | 2012-2019 | 2 104 | 263 |
|  | 2020-2024 | 1 895 | 379 |
| Other malaise and fatigue (ICD9=78079) | 2012-2019 | 204 169 | 25 521 |
|  | 2020-2024 | 197 884 | 39 577 |
| Memory Loss (ICD9=78093) | 2012-2019 | 2 357 | 295 |
|  | 2020-2024 | 1 685 | 337 |
| Generalised pain (ICD9=78096) | 2012-2019 | 2 165 | 271 |
|  | 2020-2024 | 69 080 | 13 816 |
| Altered mental status (ICD9=78097) | 2012-2019 | 1 087 | 136 |
|  | 2020-2024 | 1 677 | 335 |
| Other general symptoms (ICD9=78099) | 2012-2019 | 52 410 | 6 551 |
|  | 2020-2024 | 97 440 | 19 488 |
| Gait abnormality (ICD9=7812) | 2012-2019 | 168 359 | 21 045 |
|  | 2020-2024 | 94 484 | 18 897 |
| Lack of coordination (ICD9=7813) | 2012-2019 | 9 975 | 1 247 |
|  | 2020-2024 | 5 813 | 1 163 |
| Shortness of breath (ICD9=78605) | 2012-2019 | 7 404 | 926 |
|  | 2020-2024 | 9 472 | 1 894 |
| Cough (ICD9=7862) | 2012-2019 | 29 081 | 3 635 |
|  | 2020-2024 | 46 318 | 9 264 |

Table S19: Long COVID-19 symptoms diagnosis in the pre- and post-pandemic periods. Period (pre or post pandemic), number of events, number of events per year.

### Sensitivity analyses

#### Basic and full adjustment survival analysis comparison

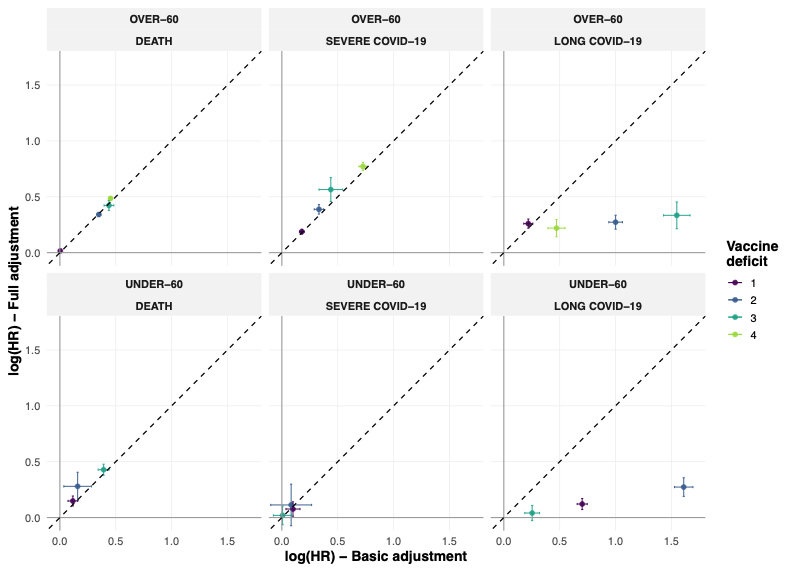

Figure S8: Comparison of undervaccination levels exposure effects (log hazard ratio) between basic adjustment and full adjustment analysis. Adjustments were included for: age group, sex, continent of birth, urbanisation classification, comorbidity score, regional healthcare provider, number of COVID-19 PCR tests in the last six months, number of positive COVID-19 PCR tests in the previous six months, last positive COVID-19 PCR test, COVID-19 hospitalisation, and non-COVID-19 hospitalisation.

#### Long COVID-19 definitions sensitivity

We conducted a sensitivity analysis for the definition of the long COVID-19 outcome, removing the necessity of a COVID-19 positive PCR test one month before the event to classify it as long COVID-19, with the hypothesis of undertesting in the later stages of the pandemic until December 2024. Surprisingly, in the under-60 analysis, the three-vaccination deficit was associated with lower odds of long COVID-19 symptoms (HR = 0.879 [0.845-0.91]), while the remaining vaccine deficits showed a lower HR than the main analysis, in line with a less restrictive long COVID-19 specific definition.

Infection ascertainment (main vs sensitivity):

- Main definition: infection required a positive SARS-CoV-2 PCR test, and symptoms had to occur ≥1 month after that PCR-positive date.
- Sensitivity definition: we removed the requirement for PCR-confirmed infection to account for possible under-testing in later pandemic phases.

| **Vaccine deficit** | **Full adjustment HR** | **P-value** | **Full adjustment 2.5%**  **Confidence limit** | **Full adjustment 97.5%**  **Confidence limit** |
| --- | --- | --- | --- | --- |
| 1 | 1·15 | <0·000 | 1·14 | 1·17 |
| 2 | 1·28 | <0·000 | 1·25 | 1·32 |
| 3 | 1·37 | <0·000 | 1·26 | 1·48 |
| 4 | 1·05 | 0·003 | 1·02 | 1·09 |

Table S20: Long COVID-19 Sensitivity analysis - Over 60

| **Vaccine deficit** | **Full adjustment HR** | **P-value** | **Full adjustment 2.5%**  **Confidence limit** | **Full adjustment 97.5%**  **Confidence limit** |
| --- | --- | --- | --- | --- |
| 1 | 1·16 | <0·000 | 1·137 | 1·19 |
| 2 | 1·33 | <0·000 | 1·25 | 1·42 |
| 3 | 0·88 | <0·000 | 0·85 | 0·91 |

Table S21: Long COVID-19 Sensitivity analysis - Under 60

Counterfactual simulations
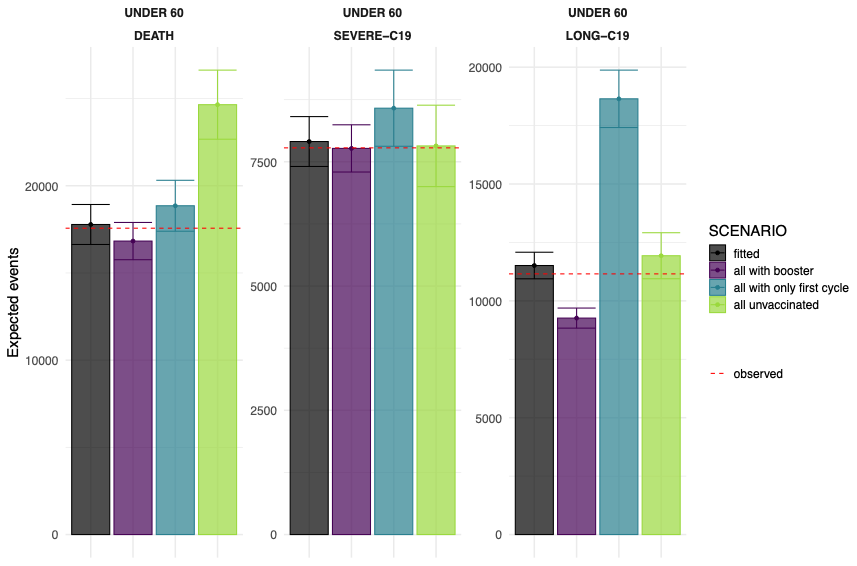

Figure S9: Fitted, observed and simulated number of cases with 95% confidence intervals of the scenarios in the under60 subgroup

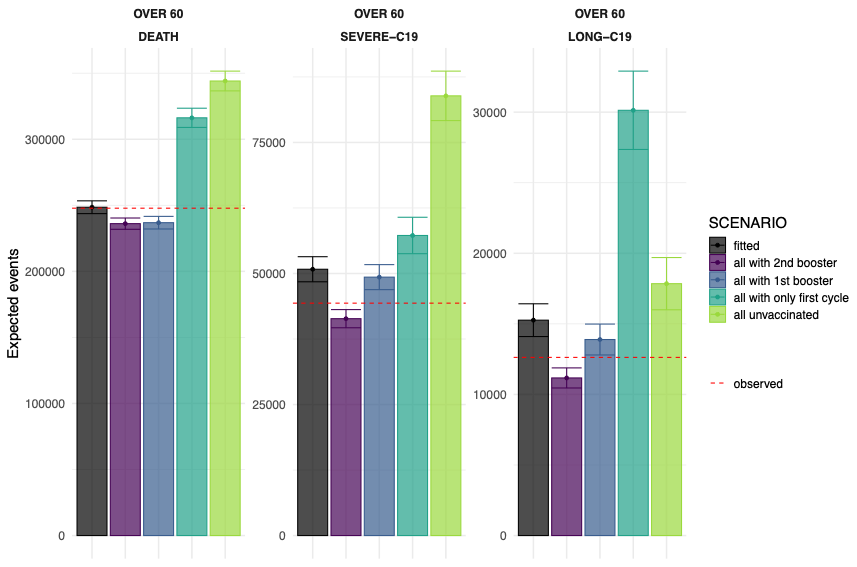

Figure S10: Fitted, observed and simulated number of cases with 95% confidence intervals of the scenarios in the over60 subgroup

| **Observed** | **Expected** | **Upper 95%**  **confidence bound** | **Lower 95%**  **confidence bound** | **Difference** | **Scenario** | **Outcome** |
| --- | --- | --- | --- | --- | --- | --- |
| 17 566 | 17 779 | 16 633 | 18 925 | 213 | Fitted | Death |
| 17 566 | 24 649 | 22 667 | 26 630 | 7 083 | All unvaccinated | Death |
| 17 566 | 18 853 | 17 392 | 20 313 | 1 287 | All with only first cycle | Death |
| 17 566 | 16 827 | 15 759 | 17 896 | -739 | All with booster | Death |
| 7 780 | 7 907 | 7 406 | 8 409 | 127 | Fitted | Severe-C19 |
| 7 780 | 7 819 | 7 000 | 8 638 | 39 | All unvaccinated | Severe-C19 |
| 7 780 | 8 578 | 7 812 | 9 344 | 798 | All with only first cycle | Severe-C19 |
| 7 780 | 7 769 | 7 294 | 8 244 | -11 | All with booster | Severe-C19 |
| 11 155 | 11 513 | 10 944 | 12 081 | 358 | Fitted | Long-C19 |
| 11 155 | 11 931 | 10 947 | 12 916 | 776 | All unvaccinated | Long-C19 |
| 11 155 | 18 646 | 17 417 | 19 874 | 7 491 | All with only first cycle | Long-C19 |
| 11 155 | 9 263 | 8 834 | 9 692 | -1 892 | All with booster | Long-C19 |

Table S19: Simulation results - Under 60. Observed and expected (from basic adjustment model) number of cases, upper and lower 95% confidence bounds and difference between observed and expected cases for each outcome and scenario.

| **Observed** | **Expected** | **Upper 95%**  **confidence bound** | **Lower 95%**  **confidence bound** | **Difference** | **Scenario** | **Outcome** |
| --- | --- | --- | --- | --- | --- | --- |
| 247 817 | 248 539 | 243 701 | 253 376 | 722 | Fitted | Death |
| 247 817 | 344 093 | 336 611 | 351 576 | 96 276 | All unvaccinated | Death |
| 247 817 | 316 221 | 308 945 | 323 496 | 68 404 | All with only first cycle | Death |
| 247 817 | 236 846 | 232 067 | 241 625 | -10 971 | All with 1st booster | Death |
| 247 817 | 236 106 | 231 834 | 240 379 | -11 711 | All with 2nd booster | Death |
| 44 341 | 50 808 | 48 416 | 53 200 | 6 467 | Fitted | Severe-C19 |
| 44 341 | 83 888 | 79 169 | 88 607 | 39 547 | All unvaccinated | Severe-C19 |
| 44 341 | 57 252 | 53 777 | 60 727 | 12 911 | All with only first cycle | Severe-C19 |
| 44 341 | 49 302 | 46 916 | 51 689 | 4 961 | All with 1st booster | Severe-C19 |
| 44 341 | 41 377 | 39 643 | 43 110 | -2 964 | All with 2nd booster | Severe-C19 |
| 12 625 | 15 261 | 14 101 | 16 422 | 2 636 | Fitted | Long-C19 |
| 12 625 | 17 848 | 15 998 | 19 698 | 5 223 | All unvaccinated | Long-C19 |
| 12 625 | 30 131 | 27 355 | 32 907 | 17 506 | All with only first cycle | Long-C19 |
| 12 625 | 13 891 | 12 796 | 14 986 | 1 266 | All with 1st booster | Long-C19 |
| 12 625 | 11 168 | 10 457 | 11 879 | -1 457 | All with 2nd booster | Long-C19 |

Table S20: Simulation results - Over 60. Observed and expected (from basic adjustment model) number of cases, upper and lower 95% confidence bounds and difference between observed and expected cases for each outcome and scenario.
